# Exploiting multivariate network meta-analysis: A calibrated Bayesian composite likelihood inference

**DOI:** 10.1101/2024.06.25.24309477

**Authors:** Yifei Wang, Lifeng Lin, Yu-Lun Liu

## Abstract

Multivariate network meta-analysis has emerged as a powerful tool in evidence synthesis by incorporating multiple outcomes and treatments. Despite its advantages, this method comes with methodological challenges, such as the issue of unreported within-study correlations among treatments and outcomes, which potentially lead to misleading conclusions. In this paper, we proposed a calibrated Bayesian composite likelihood approach to overcome this limitation. The proposed method eliminated the need to specify a full likelihood function while allowing for the unavailability of within-study correlations among treatments and outcomes. Additionally, we developed a hybrid Gibbs sampler algorithm along with the Open-Faced Sandwich post-sampling adjustment to enable robust posterior inference. Through comprehensive simulation studies, we demonstrated that the proposed approach yielded unbiased estimates while maintaining coverage probabilities close to the nominal level. Furthermore, we implemented the proposed method on two real-world network meta-analysis datasets; one comparing treatment procedures for the root coverage and another comparing treatments for anaemia in chronic kidney disease patients.

## 1. Introduction

Network meta-analysis (NMA), also known as multiple treatments meta-analysis or mixed treatment comparisons, has emerged as a pivotal tool for synthesizing the effectiveness and safety of multiple treatment regimens in medical and healthcare domains (Lumley, 2002; Lu and Ades, 2006; White et al., 2012; Salanti, 2012; Riley et al., 2017). This is evident by the growing number of NMAs, as a recent comprehensive review by Petropoulou et al. (2017) identified 456 NMAs from randomized trials across different domains. The rationale behind NMA lies in estimating pooled treatment effects across at least three interventions by combining both direct and indirect evidence into a coherent framework. This approach overcomes the limitations of scarce head-to-head trials and allows for borrowing indirect evidence through a common comparator. The NMA enables the establishment of a treatment hierarchy, ranking candidate interventions from most to least effective (Cipriani et al., 2013). Numerous approaches for treatment ranking have been developed and widely adopted in healthcare-related research, such as the surface under the cumulative ranking curve (SUCRA) (Salanti et al., 2011), P-score (Rücker and Schwarzer, 2015), among others.

While NMAs typically focus on single outcomes, real-world scenarios often require evaluating multiple outcomes simultaneously, such as benefit-risk assessment and health economic evaluation. Since these outcomes of interest are measured within the same population in the same study, they are likely to be correlated. As suggested by Jackson et al. (2011), correlations among multiple outcomes are potentially informative and worth borrowing of strength. Multivariate NMAs allow for the incorporation of correlation estimations, in which correlations between multiple outcomes can occur at both the between-study and within-study levels (Riley et al., 2017). However, obtaining within-study correlations poses a challenge due to rarely reporting in trials (Riley, 2009). Ignoring the possibility of within-study correlations tends to distort estimates of relative treatment effects, leading to increased mean-square errors and standard errors of pooled effect estimates (Riley, 2009). Standard multivariate NMAs or meta-analyses typically assume either known or zero within-study correlations. Alternatively, assumed correlation coefficients, possibly combined with sensitivity analysis (Waddingham et al., 2020), can be used to achieve within-study correlations. Riley et al. (2008) proposed a single overall correlation parameter by combining within-study and between-study correlations in the multivariate meta-analysis framework. Furthermore, Wei and Higgins (2013) derived approximate formulations for assessing within-study correlations across various pairs of treatment effect measurements.

Recent advancements in the methodologies of multivariate NMAs have given rise in the literature. Efthimiou et al. (2014) introduced a joint model of two correlated dichotomous outcomes in a network and elicited expert opinions to inform within-study correlations. Expanding on their work, Efthimiou et al. (2015) further incorporated both within-study and between-study correlations into a Bayesian framework. They also implemented the overall correlations proposed by Riley et al. (2008) as an alternative model to tackle the absence of within-study correlations in multivariate NMAs. Hong et al. (2016) presented an alternative Bayesian framework, building on the work of Lu and Ades (2004), for modeling multiple correlated outcomes; however, their proposed approach did not account for within-study correlations. Jackson et al. (2018) proposed a matrix-based method of moment technique for modeling multivariate NMAs, allowing for between-study heterogeneity and inconsistency. Furthermore, Liu et al. (2018) devised a Bayesian hierarchical approach based on the bivariate Clayton copula model, which leveraged information across multiple correlated outcomes and treatments to partially mitigate the impact of outcome reporting bias. A recent contribution by Duan et al. (2023) introduced personalized treatment ranking procedures which enabled the consideration of individuals’ preferences across multiple outcomes.

In this paper, we proposed a Bayesian composite likelihood-based method for multivariate NMAs to address two potential challenges: the unavailability of within-study correlations and computational burden. While composite likelihood methods (Lindsay, 1988; Varin et al., 2011) are common in spatial extremes research (Pauli et al., 2011; Ribatet et al., 2012), it has not been extensively explored in the context of multivariate NMAs. The proposed method offers several advantages over existing Bayesian methods for multivariate NMAs. First, the proposed method ensured valid statistical inference without requiring the knowledge of within-study correlations among outcomes and treatments. This can be achieved by calibrating the composite likelihood to approximate the posterior distribution of parameters using the Markov chain Monte Carlo algorithm. Second, it is acknowledged that the posterior distribution derived from a composite likelihood method may lead to overly precise estimates (Pauli et al., 2011; Ribatet et al., 2012). Adjusting composite likelihoods in the Bayesian framework is common, but post-calibration may lead to overestimation and necessitate additional computational procedures. We tackled this issue by employing the Open-Faced Sandwich adjustment on posterior samples to ensure accurate asymptotic variance and preserve the proper shapes of posterior distributions. Finally, the proposed method achieved significant computational efficiency gains compared to existing Bayesian methods through the implementation of a parallel sampling strategy.

The remainder of this paper is organized as follows. In Section 2, we present two motivating NMA examples. In Section 3, we formalize the Bayesian composite likelihood approach for multivariate NMAs using the Markov chain Monte Carlo algorithm. We introduce calibration on posterior samples via the Open-Faced Sandwich adjustment. In Section 4, we evaluate the performance and statistical properties of the proposed method by using simulation studies across various scenarios. In Section 5, we demonstrate the application of the proposed method with two motivating NMA examples. Finally, Section 6 concludes the paper with a discussion and provides recommendations for future research.

## 2. Motivating examples

### 2.1 Comparison of coronally advanced flap-based root coverage procedures

Several procedures combined with the coronally advanced flap (CAF) have been proposed to treat gingival recessions. Cairo et al. (2008) showed that using a connective tissue graft (CTG) or enamel matrix derivative (EMD) in conjunction with CAF increases the probability of obtaining complete root coverage (CRC) in Miller Class I and II single gingival recessions. Cheng et al. (2007) found that the clinical outcomes of CAF were more predictable when the EMD procedure was added. However, these systematic reviews did not compare all possible treatment procedures. To address this gap, Buti et al. (2013) employed a Bayesian network meta-analysis (Lu and Ades, 2006) to compare and rank the efficacy of seven CAF-based treatments, including CAF, CAF plus connective tissue graft (CAF+CTG), CAF plus enamel matrix derivative (CAF+EMD), CAF plus barrier membrane (CAF+BM), CAF plus acellular dermal matrix (CAF+ADM), CAF plus platelet rich plasma (CAF+PRP), CAF plus collagen matrix (CAF+CM), and CAF plus human fibroblast derived dermal substitute (CAF+HF-DDS). The study included 29 trials with a focus on four clinical outcomes: recession reduction (RecRed), clinical attachment gain (CALgain), keratinized tissue gain (KTgain), and complete root coverage (CRC). However, the within-study correlations between treatments and outcomes were not reported in this NMA. The evidence networks are illustrated in Figure S1 of Appendix A in the Supplementary Materials. Consequently, they concluded that CAF combined with CTG represented the most efficacious treatment procedure.

### 2.2 Comparison of treatments for anaemia in adults with chronic kidney disease

Anaemia, characterized by a deficiency in red blood cells, is a frequent complication among patients with chronic kidney disease (CKD). This condition can manifest with symptoms such as fatigue and shortness of breath and, in severe cases, may necessitate blood transfusions. Erythropoiesis-stimulating agents (ESAs) are commonly administrated to CKD patients to treat anaemia; however, recent studies suggested potential cardiovascular risks associated with ESA use (Hanna et al., 2021). To further examine the efficacy and safety of various ESAs, Chung et al. (2023) conducted an NMA from 74 published trials up to April 29, 2022. This analysis compared six ESA treatments (including epoetin alfa, epoetin beta, darbepoetin alfa, methoxy polyethylene glycol-epoetin beta, biosimilar epoetin, and biosimilar darbepoetin) across 9 clinically relevant outcomes, such as blood transfusion, cardiovascular mortality, and kidney failure, etc. The networks of evidence are illustrated in Figure S2 of Appendix A in the Supplementary Materials. They employed a multivariate meta-regression method for NMAs, as proposed by White et al. (2012), assuming a common heterogeneity variance structure and known within-study correlations. Consequently, Chung et al. (2023) concluded that the relative efficacy of different ESAs remained unclear.

## 3. Methodology

### 3.1 Notation and model specifications

Suppose that *I* studies (*i* = 1*, … , I*) have been conducted to compare a total of 𝒯 treatments denoted by 𝒯 = {*A, B, C*, … }. Each study compares at least two treatments in 𝒯. Let *d* = 1*, … , D* index the designs or sets of treatments compared within a study. Define *n_d_* as the total number of studies for the *d*th design, each involving a comparison of *T_d_* treatments. For example, if *d* = 1 corresponds to a three-arm design where treatments *A*, *B*, and *C* are compared (thus 𝒯_1_ = 3), and there is only one study following this three-arm design, then *n*_1_ = 1. Assume that a set of *L* outcomes is of interest, where *L* continuous outcomes consist of standardized mean differences or other transformed continuous scales, such as log odds ratios or log risk ratios. Let 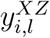 be the observed contrast of treatment *Z* (*Z* ∈ 𝒯) with treatment *X* (*X* ∈ 𝒯) for the *i*th study and the *l*th outcome (*l* = 1*, … , L*), along with a standard error 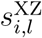.

For simplicity of illustration and without loss of generality, we consider a network involving three treatments with two distinct yet potentially correlated continuous outcomes, denoted by 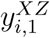 and 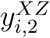, where *Z, X* ∈ 𝒯 = *{A, B, C}*. Any treatment can be chosen as a reference; for the sake of simplicity, treatment *A* is chosen as a reference throughout this section. When comparing the treatments *A*, *B* and *C*, possible designs include two-arm designs *AB*, *BC*, *AC*, as well as a three-arm design *ABC*. Let 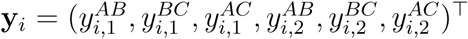 and 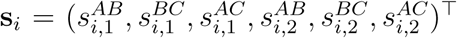 represent the observed treatment effects and their associated standard errors for the *i*th study, respectively. The random-effects model for the observed data is expressed as follows,

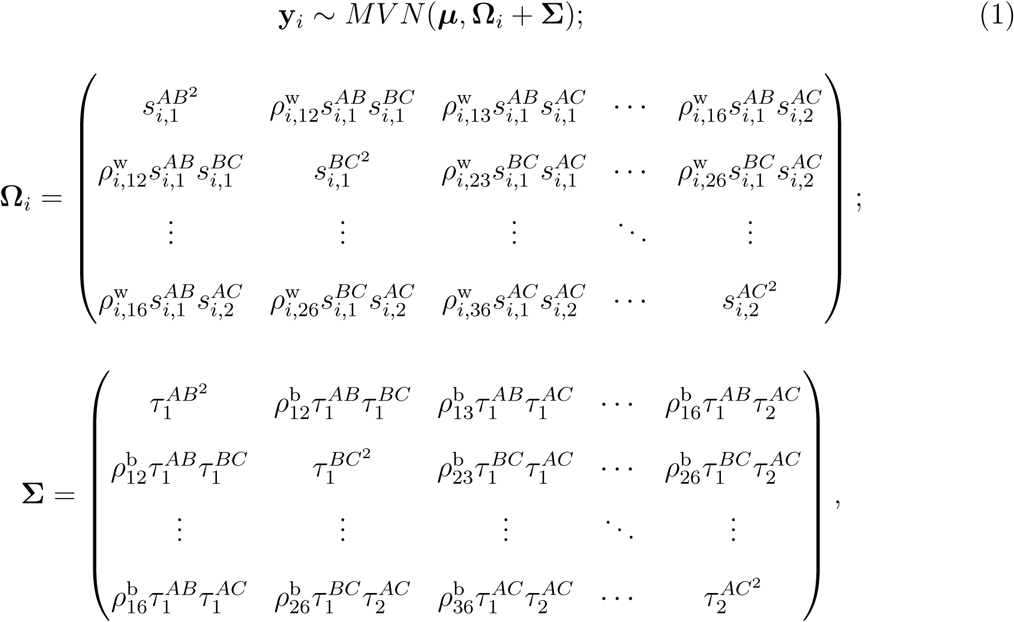

where 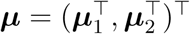, and 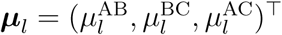 represents the overall means of estimates. **Ω***_i_* is the 6 *×* 6 within-study variance-covariance matrix;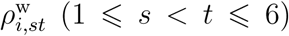 captures the within-study correlations, but its estimation is often challenged by limited available data (Riley et al., 2008; Riley, 2009; Kirkham et al., 2012). We denote the within-study correlation matrix for each study as 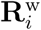. Furthermore, **Σ** is the 6 *×* 6 between-study variance-covariance matrix; 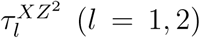 and 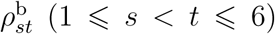 describe the between-study heterogeneity and correlations. The between-study heterogeneity variances are denoted by 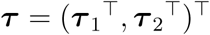 with 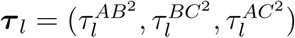 for *l* = 1, 2, and the between-study correlation matrix is represented as **R**^b^.

### 3.2 Proposed method

The full likelihood function presented in Equation (1) often encounters computational challenges due to the high dimensionality of the parameter space represented by 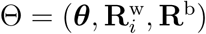. This space includes parameters of primary interest ***θ*** = (***µ***, ***τ***) and nuisance parameters 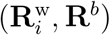. This complexity motivates the exploration of computationally attractive approximations under a Bayesian framework, such as the composite likelihood function. More specifically, this strategy is to replace the full likelihood function with a surrogate composite likelihood (Lindsay, 1988; Varin et al., 2011), which shares similar properties to the standard full likelihood. The concept behind the composite likelihood paradigm (Lindsay, 1988) lies in constructing a set of pseudo-likelihood functions based on either conditional or marginal likelihood components. Let 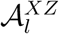 be a set of studies containing effect sizes along with their associated standard errors for outcome *l*, with a focus on comparing treatments *X* and *Z*. It is worth noting that studies in each 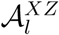 are likely to overlap due to the inclusion of multiple outcomes and treatment comparisons in single studies. Considering all the 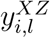 for studies in 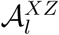, the log composite likelihood function for the observed data becomes

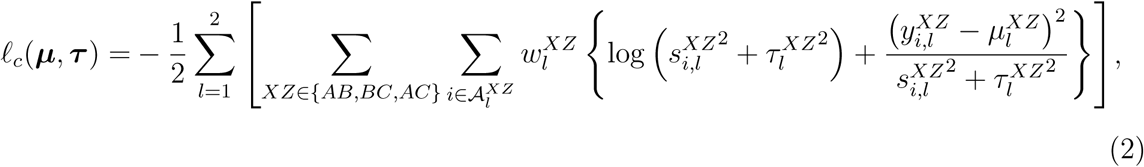

where 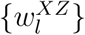 represents non-negative weights. We set 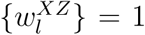 for all *XZ*, followed by post-sample adjustment that implicitly incorporates different weights.

#### 3.2.1 Prior and posterior distributions

In the log composite likelihood function for observed data (as shown in Equation (2)), the parameters to be estimated consist of the treatment effect 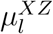 and the between-study variance 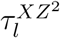 for the outcome *l* between treatments *X* and *Z*. Assuming no prior knowledge, we assign non-informative prior distributions to these unknown parameters. For the outcome *l* between treatments *X* and *Z*, we choose a normal prior distribution *N* (0, 10000) for the treatment effect 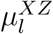 and an inverse-gamma distribution *IG*(0.001, 0.001) for the between-study variance 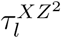.

To generate samples of the parameters of interest from their posterior distributions, we leverage the observed data to derive posterior kernel functions from the composite likelihood function and chosen priors. Given the observed data **y** and the vector of between-study variance ***τ***, the posterior kernel function for the treatment effect 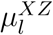 is given by

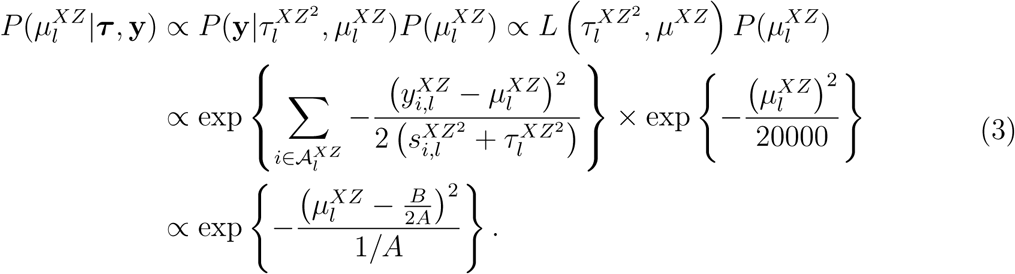

Thus, the posterior distribution, 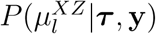, follows a normal distribution, that is,

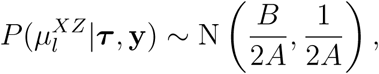

where

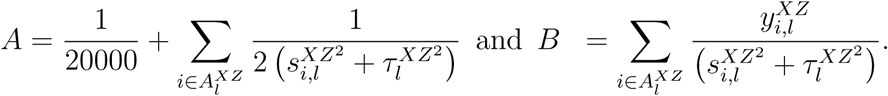

Similarly, conditioning on the observed data **y** and the treatment effect ***µ***, the posterior kernel function for the between-study variance 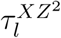 is

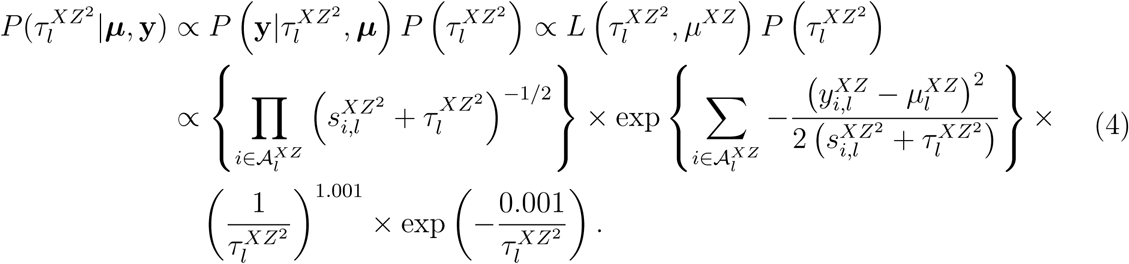

Note that the above posterior kernel function does not align with any known distributions, resulting in non-conjugate posterior distributions. To tackle this challenge, we tailor the Metropolis–Hastings (MH) step within the Gibbs sampler to sample from the posterior distribution of the between-study variance 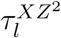 in the following subsection. The detailed derivation can be found in Appendix B of the Supplementary Materials.

#### 3.2.2 Markov chain Monte Carlo algorithm: a hybrid of Gibbs samplers and Metropolis–Hastings

To approximate the posterior distributions of model parameters, we utilize the Markov chain Monte Carlo (MCMC) algorithm. This framework enables efficient approximation of complex posterior distributions by constructing a sequence of random observations. Within the MCMC, we propose a Gibbs sampler, specifically designed to exploit the conditional dependence between model parameters. The iterative sampler operates by sequentially drawing samples from the composite posterior distribution of each parameter in turn, conditioned on the current values of all other parameters and the observed data, as illustrated in **Algorithm 1**.

##### Algorithm 1 Gibbs sampler algorithm

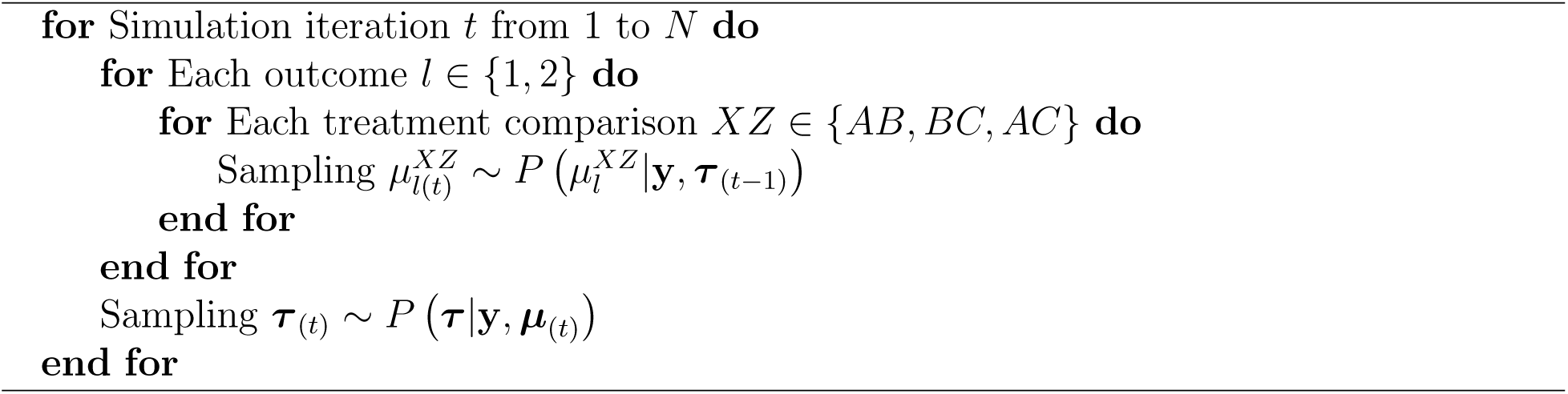

Each iteration of the hybrid Gibbs sampler employs a combination of Gibbs and MH steps to overcome the lack of a closed-form solution for the full conditional distribution of between-study variance 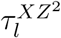. For each iteration, we leverage a truncated normal distribution conditioned on the values of between-study variance obtained in the previous iteration. However, as the number of treatments increases, the computational cost can grow exponentially due to growing number of between-study variances. To mitigate this, we exploit the posterior kernel function, as shown in Equation (4), in conjunction with the proposed truncated normal distribution to group the between-study variances 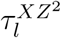 into blocks and sample them together. The implementation details of the MH step are provided in **Algorithm 2**.

##### Algorithm 2 Metropolis–Hastings step for *τ*

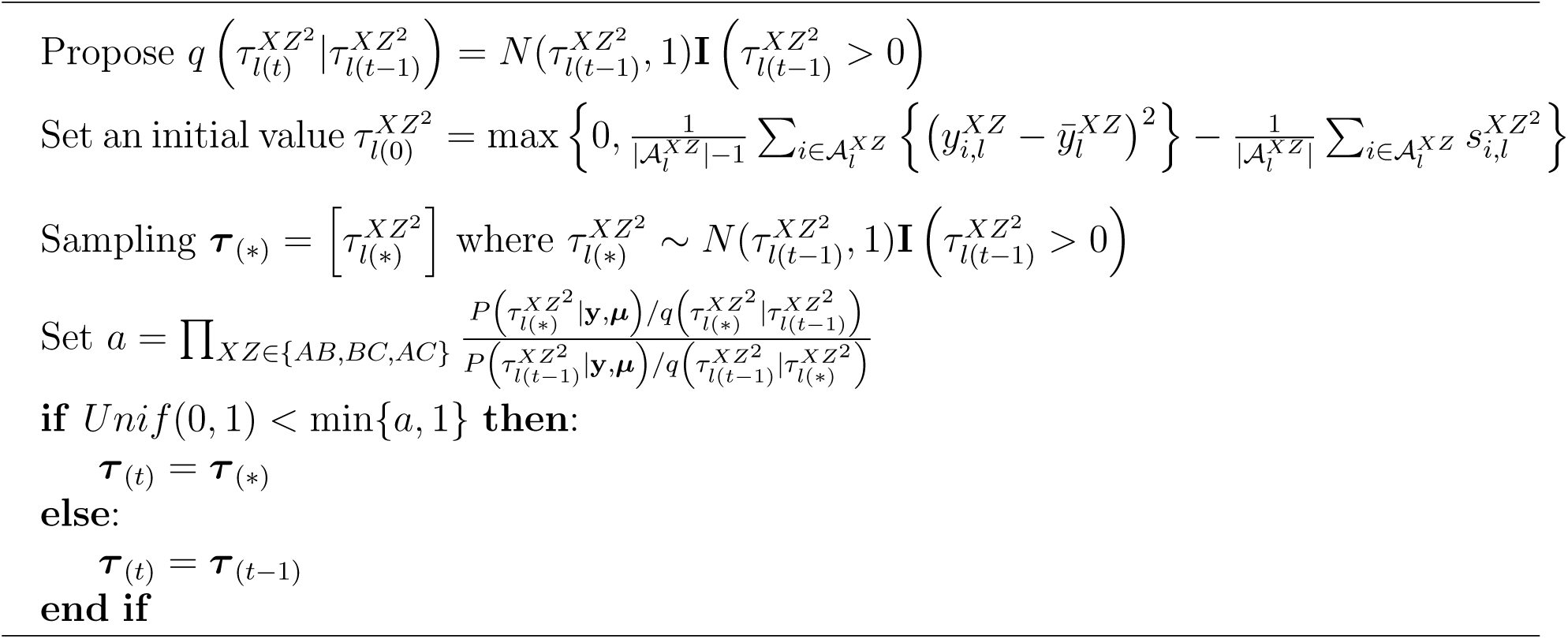

We sample each individual treatment sequentially. Since the posterior distribution for treatment effect follows a normal distribution, we directly sample each treatment effect based on other parameters obtained in the previous step. Details of the Gibbs step are provided in **Algorithm 3**.

##### Algorithm 3 Gibbs step for *µ*

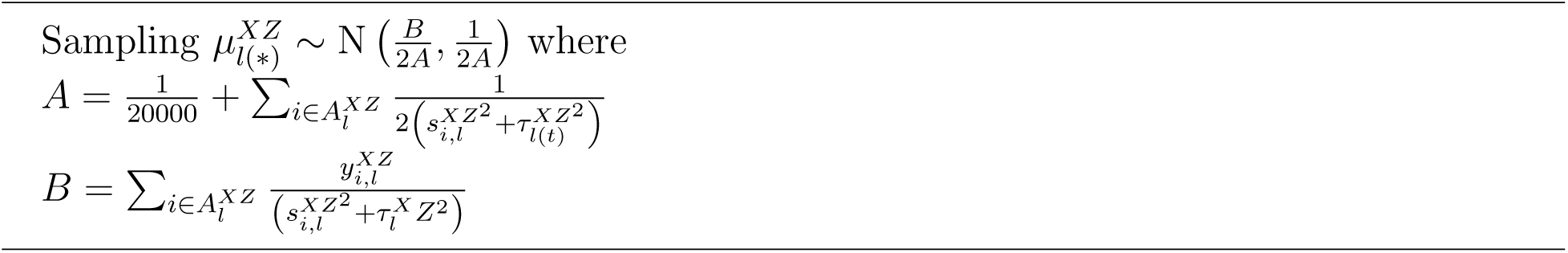

#### 3.2.3 Posterior inference with the Open-Faced Sandwich adjustment

Our proposed approach involves substituting the full likelihood function with the composite likelihood function within the posterior density to generate the posterior samples. However, it is well-acknowledged that the posterior distribution derived from a composite likelihood can lead to overly precise estimates (Pauli et al., 2011; Ribatet et al., 2012). Thus, we employ the Open-Faced Sandwich (OFS) adjustment on the posterior samples to ensure asymptotically valid parameter estimation. This method, building upon the sandwich-type estimators, applies to an objective function involving both the observed data and unknown parameters, yielding asymptotically valid estimations (Shaby, 2014). As our initial composite posterior estimates 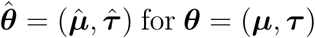 correspond to the maximum a posteriori (MAP) estimates, we then construct the log composite posterior as the objective function by summing the log-composite likelihood in Equation (2) and the logarithm of the prior distributions

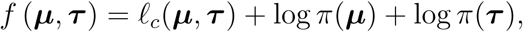

where *π*(***µ***) and *π*(***τ***) denote the respective prior distributions for ***µ*** and ***τ***. Following the principles of the OFS adjustment, we compute the score vector and Hessian matrix at the initial composite posterior estimates, 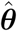, using the Newton–Raphson method

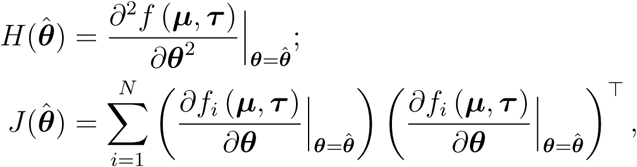

where *f_i_* (***µ***, ***τ***) is the objective function given the data of study *i*. While various numerical methods are feasible, we opt for the Newton-Raphson method due to its flexibility in handling scenarios with varying numbers of treatments and outcomes. Upon obtaining the score vector and Hessian matrix, we can derive the expression for the adjusted composite posterior

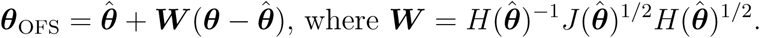

Importantly, the initial MAP estimate remains unaffected by the OFS adjustment. Compared with the in-sampling adjustment methods such as Moment-type adjustment (Chandler and Bate, 2007), the OFS method offers two key advantages. First, by performing the post-sampling adjustments, the OFS method eliminates the requirement to calculate the maximum likelihood estimate at each sampling step, thereby significantly improving computational efficiency. Second, unlike the in-sampling methods that modify individual samples, the OFS method avoids distorting the sampling process from its stationary distribution.

#### 3.2.4 Parallel sampling for computational efficiency gains

By incorporating the composite likelihood function into the posterior distribution, we can efficiently generate independent posterior samples for each outcome. This allows us to leverage a parallel sampling strategy using the R packages **doParallel** and **foreach**. Specifically, we simultaneously sample from the posterior distributions of all outcomes. Upon completion of the sampling procedure, we combine the posterior samples across different outcomes and apply the OFS adjustment. This parallel sampling strategy enhances the efficiency of the sampling process, particularly when dealing with multiple outcomes. The execution time comparisons are illustrated in the next simulation section.

## 4. Simulation study

### 4.1 Data generating mechanisms

For the simulation study, we employed a contrast-based multivariate NMA with a three-arm design (i.e., *A*, *B*, and *C*, with *A* as a reference) for two continuous outcomes of primary interest. Of note, ratio measures such as odds ratios and risk ratios, along with their log transformations, can be treated as continuous variables. Within the simulated network, consistency was assumed to be satisfied, implying 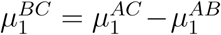 and 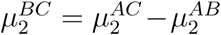. Data were drawn from a multivariate normal distribution following Equation (1), with specified treatment effects: 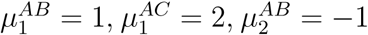, and 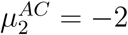. We assessed the performance of the proposed method without and with the OFS adjustment (i.e., BCL and BCL-OFS), and the Bayesian full-likelihood method (i.e., BFL) across various scenarios, including (i) equality of between-study heterogeneity variance (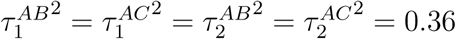 under common between-study heterogeneity variance; 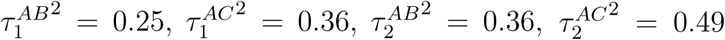 under unequal between-study heterogeneity variance), (ii) varying numbers of studies (*I* = 5, 10, 20, 40, and 100), and (iii) various within-study and between-study correlation structures. Specifically, we evaluated the magnitude of correlations, from low (*ρ^w^* = *ρ^b^* = 0.2), moderate (*ρ^w^* = *ρ^b^* = 0.5), to high (*ρ^w^*= *ρ^b^* = 0.8), and varying correlations

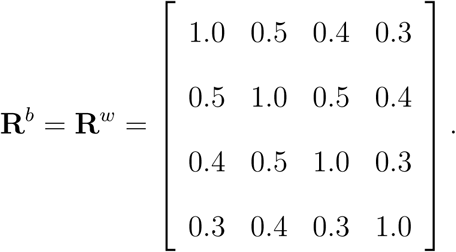

For each scenario, 1, 000 NMA datasets were generated. Detailed specifications of the simulation design are provided in Appendix C of the Supplementary Materials.

To evaluate the computational efficiency of the BCL method, we compared its execution time against the BFL methods using the simulated data. The BFL methods employed the full-likelihood function as described in Equation (1), assuming a moderate within-study correlation of *ρ^w^*= 0.5. Both BCL and BFL methods utilized the same prior distributions. The Markov Chain length was run for 3, 000 iterations for every parameter, discarding the first 1, 500 as burn-in and collecting posterior samples every 5th iteration afterward.

### 4.2 Simulation results

We evaluated the performance of BCL, BCL-OFS, and BFL methods in terms of bias and coverage probability of 95% credible intervals (CrI) for the estimated treatment effects. Table 1 presents the bias of treatment effect estimates for both outcomes when data were generated under a common between-study heterogeneity variance. Of note, the OFS adjustment did not alter the location of the initial composite likelihood distribution; in other words, treatment effect estimates remained consistent regardless of the OFS adjustment. As shown in Table 1, the BCL method overall yielded nearly unbiased pooled estimates for treatment comparisons *AB* and *AC* across most scenarios and both outcomes. The magnitude of high within-study correlations did not noticeably impact the bias of treatment effects. However, scenarios with a limited number of studies (*I* = 5 or 10), particularly those with moderate to varying within-study correlations, exhibited slightly larger bias. Furthermore, an increasing number of studies led to reduced bias for both outcomes, especially in scenarios with varying within-study correlations. Similar patterns were observed in scenarios with unequal between-study heterogeneity variance (Tables S1 and S2 of the Supplementary Materials).

**Table 1.** Summary of 1, 000 simulations with I = 5, 10, 20, 40 and 100: bias and coverage probability (CP) of pooled estimates of AB and AC treatment comparisons for two outcomes. The data generating mechanism was through a common between-study heterogeneity variance with 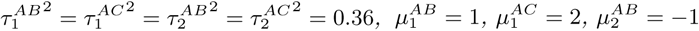 and 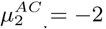, as well as high correlations ρ^w^ = ρ^b^ = 0.8, moderate correlations ρ^w^ = ρ^b^ = 0.5, low correlations ρ^w^ = ρ^b^ = 0.2 and varying correlations. Results were based on the proposed BCL method.

|  | High Correlation |  |  |  | Moderate Correlation |  |  |  | Low Correlation |  |  |  | Varying Correlation |  |  |  |
| --- | --- | --- | --- | --- | --- | --- | --- | --- | --- | --- | --- | --- | --- | --- | --- | --- |
| | $\mu_1^{AB}$ | $\mu_1^{AC}$ | $\mu_2^{AB}$ | $\mu_2^{AC}$ | $\mu_1^{AB}$ | $\mu_1^{AC}$ | $\mu_2^{AB}$ | $\mu_2^{AC}$ | $\mu_1^{AB}$ | $\mu_1^{AC}$ | $\mu_2^{AB}$ | $\mu_2^{AC}$ | $\mu_1^{AB}$ | $\mu_1^{AC}$ | $\mu_2^{AB}$ | $\mu_2^{AC}$ |
| **I=5** |  |  |  |  |  |  |  |  |  |  |  |  |  |  |  |  |
| Bias | 0.008 | 0.001 | 0.007 | 0.002 | 0.006 | 0.008 | 0.011 | 0.008 | 0.018 | 0.016 | 0.009 | 0.010 | 0.005 | 0.007 | 0.010 | 0.006 |
| CP | 0.882 | 0.882 | 0.881 | 0.884 | 0.912 | 0.907 | 0.901 | 0.924 | 0.906 | 0.908 | 0.909 | 0.921 | 0.909 | 0.895 | 0.921 | 0.912 |
| **I=10** |  |  |  |  |  |  |  |  |  |  |  |  |  |  |  |  |
| Bias | 0.009 | 0.000 | 0.005 | 0.005 | 0.003 | 0.005 | 0.007 | 0.003 | 0.009 | 0.003 | 0.005 | 0.004 | 0.006 | 0.001 | 0.013 | 0.010 |
| CP | 0.869 | 0.900 | 0.882 | 0.895 | 0.891 | 0.896 | 0.911 | 0.897 | 0.899 | 0.904 | 0.919 | 0.911 | 0.895 | 0.909 | 0.909 | 0.906 |
| **I=20** |  |  |  |  |  |  |  |  |  |  |  |  |  |  |  |  |
| Bias | 0.002 | 0.002 | 0.004 | 0.002 | 0.001 | 0.001 | 0.002 | 0.005 | 0.002 | 0.001 | 0.003 | 0.010 | 0.006 | 0.002 | 0.001 | 0.006 |
| CP | 0.886 | 0.903 | 0.899 | 0.879 | 0.904 | 0.884 | 0.898 | 0.886 | 0.910 | 0.893 | 0.906 | 0.891 | 0.899 | 0.895 | 0.916 | 0.888 |
| **I=40** |  |  |  |  |  |  |  |  |  |  |  |  |  |  |  |  |
| Bias | 0.006 | 0.009 | 0.004 | 0.006 | 0.001 | 0.002 | 0.001 | 0.005 | 0.002 | 0.001 | 0.002 | 0.001 | 0.002 | 0.005 | 0.003 | 0.002 |
| CP | 0.889 | 0.884 | 0.893 | 0.889 | 0.888 | 0.912 | 0.908 | 0.888 | 0.901 | 0.916 | 0.919 | 0.915 | 0.887 | 0.903 | 0.909 | 0.900 |
| **I=100** |  |  |  |  |  |  |  |  |  |  |  |  |  |  |  |  |
| Bias | 0.002 | 0.000 | 0.001 | 0.000 | 0.004 | 0.002 | 0.000 | 0.003 | 0.001 | 0.002 | 0.001 | 0.000 | 0.000 | 0.001 | 0.000 | 0.003 |
| CP | 0.906 | 0.907 | 0.914 | 0.912 | 0.918 | 0.910 | 0.917 | 0.922 | 0.911 | 0.922 | 0.905 | 0.915 | 0.901 | 0.897 | 0.911 | 0.906 |

Figure 1 displays the coverage probabilities of 95% CrIs for the proposed BFL and BFL-OFS methods under the scenario with a common between-study heterogeneity variance. Overall, the BCL method slightly underestimated coverage probabilities, ranging from 86.6% to 92.8% for both outcomes. This underestimation was particularly evident in scenarios with strong within-study correlations (ranging from 88.1% to 91.4%). These findings align with the previous research by Pauli et al. (2011) and Ribatet et al. (2012), suggesting that Bayesian composite likelihood methods can underestimate variance, leading to overly precise posterior distributions. Conversely, applying the OFS adjustment to the BCL method effectively mitigated this underestimation, improving coverage probabilities closer to the 95% nominal level. For scenarios with low within-study correlations, the BCL-OFS method improved coverage probabilities by 1% to 6%, achieving a range of 90.9% and 96.6%. We also observed coverage probabilities obtained from the BFL-OFS method ranged from 89.8% to 95.7% for scenarios with high within-study correlations. Similar patterns were observed when data were simulated under scenarios with unequal between-study heterogeneity variance (Figure S3 of the Supplementary Materials).

**Figure 1.**
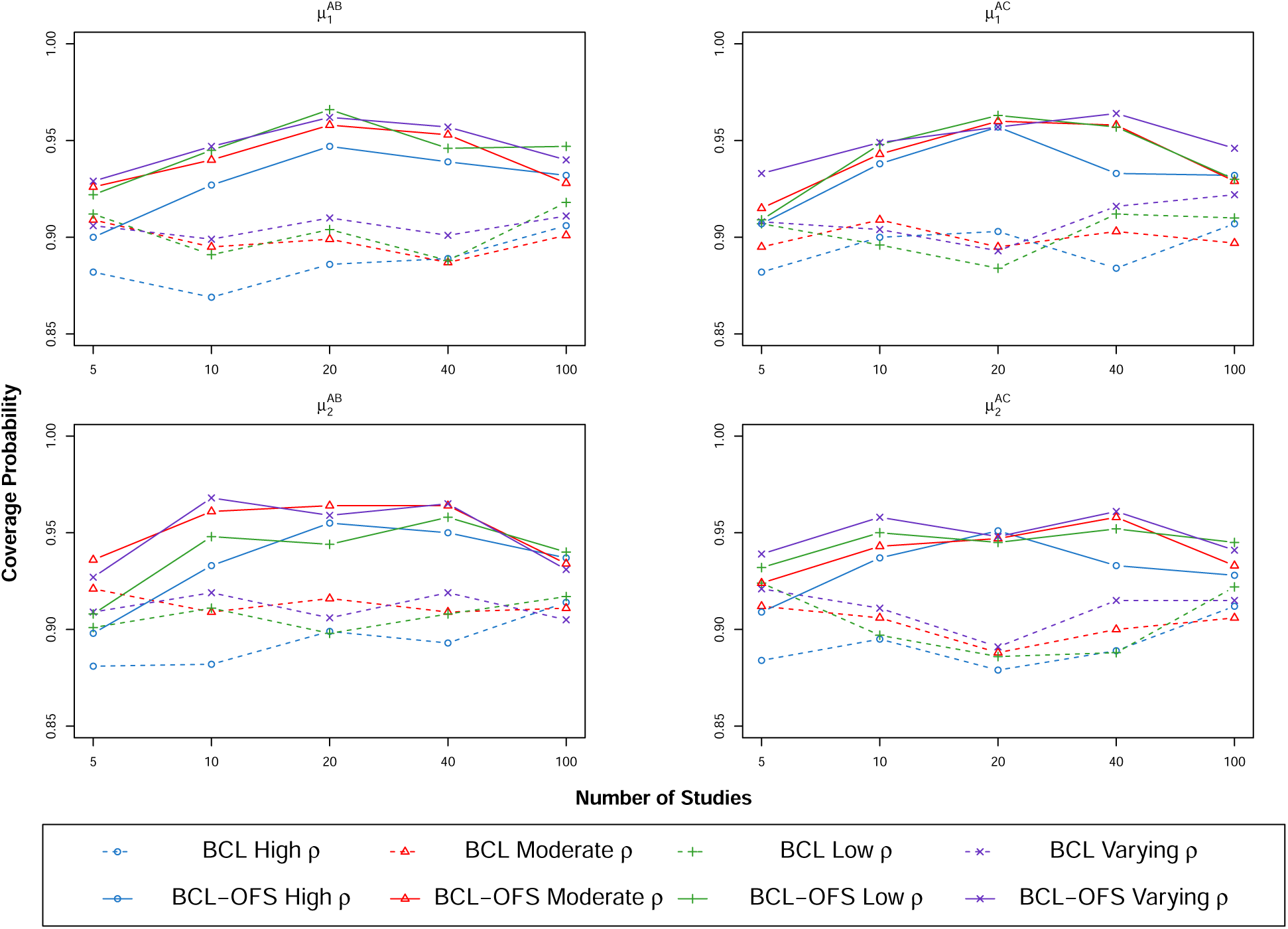
Coverage probabilities of pooled estimates for treatment comparisons *AB* and *AC* across both outcomes using the proposed BCL and BCL-OFS methods under low (*ρ^w^* = *ρ^b^* = 0.2), moderate (*ρ^w^*= *ρ^b^* = 0.5), high (*ρ^w^*= *ρ^b^* = 0.8), and varying correlations. The data generating mechanism was through a common between-study heterogeneity variance with 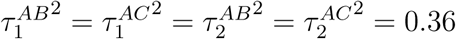.

We also compared the BCL-OFS method with the BFL method assuming moderate correlations under a common between-study heterogeneity variance, as shown in Table 2. Overall, both the BCL-OFS and BFL methods generated unbiased results. However, when the number of studies was small (e.g., *I* = 5), the BFL method tended to have underestimated coverage probabilities: 88.1% in the high correlation scenario, 85.1% in the moderate correlation scenario, 87.9% in the low correlation scenario, and 86.2% in the varying correlation scenario. The BCL-OFS method consistently had acceptable coverage probabilities above 90%. Meanwhile, we calculated the relative efficiency (RE) of the BCL-OFS method, defined as the square of the empirical standard error of the estimates from the BCL-OFL method divided by that of the BFL method. The RE ranged from 78.3% to 112.7% across all correlation structures under a common between-study heterogeneity variance. The loss of efficiency was expected because the BFL method was asymptotically the most efficient method. However, the efficiency gain of the BCL-OFS method can be attributed to the advantage of not estimating the within-study correlation parameters. As the number of studies increased, both methods exhibited coverage probabilities close to the 95% nominal level. Similar patterns were observed in scenarios with unequal between-study heterogeneity variance (Tables S3–S6 of the Supplementary Materials).

**Table 2.** Summary of 1, 000 simulations with I = 5, 10, 20, 40 and 100: bias, coverage probability (CP), and relative efficiency (RE) of pooled estimates of AB and AC treatment comparisons for two outcomes. The data generating mechanism was through a common between-study heterogeneity variance with 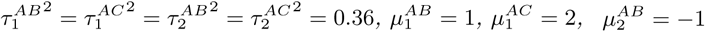, and 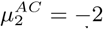, as well as high correlations ρ^w^ = ρ^b^ = 0.8, moderate correlations ρ^w^ = ρ^b^ = 0.5, low correlations ρ^w^ = ρ^b^ = 0.2, and varying correlations. Results were based on the BCL-OFS method and the BFL method assuming a moderate correlation of 0.5.

| # Study | Method | $\mu_1^{AB}$ | | | $\mu_1^{AC}$ | | | $\mu_2^{AB}$ | | | $\mu_2^{AC}$ | | |
| --- | --- | --- | --- | --- | --- | --- | --- | --- | --- | --- | --- | --- | --- |
|  |  | Bias | CP | RE | Bias | CP | RE | Bias | CP | RE | Bias | CP | RE |
| High Correlation |  |  |  |  |  |  |  |  |  |  |  |  |  |
| 5 | BFL | 0.000 | 0.898 | 1.000 | 0.003 | 0.844 | 1.000 | 0.012 | 0.911 | 1.000 | 0.004 | 0.866 | 1.000 |
|  | BCL-OFS | 0.008 | 0.900 | 1.103 | 0.001 | 0.907 | 0.922 | 0.007 | 0.898 | 1.090 | 0.002 | 0.909 | 0.999 |
| 10 | BFL | 0.002 | 0.906 | 1.000 | 0.006 | 0.849 | 1.000 | 0.005 | 0.913 | 1.000 | 0.005 | 0.854 | 1.000 |
|  | BCL-OFS | 0.009 | 0.927 | 1.084 | 0.000 | 0.938 | 0.868 | 0.005 | 0.933 | 1.028 | 0.005 | 0.937 | 0.866 |
| 20 | BFL | 0.002 | 0.942 | 1.000 | 0.001 | 0.900 | 1.000 | 0.003 | 0.941 | 1.000 | 0.003 | 0.912 | 1.000 |
|  | BCL-OFS | 0.002 | 0.947 | 1.017 | 0.002 | 0.957 | 0.898 | 0.004 | 0.955 | 0.970 | 0.002 | 0.951 | 1.024 |
| 40 | BFL | 0.001 | 0.956 | 1.000 | 0.001 | 0.929 | 1.000 | 0.003 | 0.949 | 1.000 | 0.004 | 0.936 | 1.000 |
|  | BCL-OFS | 0.006 | 0.939 | 1.031 | 0.009 | 0.933 | 0.969 | 0.004 | 0.950 | 0.985 | 0.006 | 0.933 | 1.030 |
| 100 | BFL | 0.002 | 0.954 | 1.000 | 0.001 | 0.938 | 1.000 | 0.001 | 0.950 | 1.000 | 0.002 | 0.934 | 1.000 |
|  | BCL-OFS | 0.002 | 0.932 | 0.976 | 0.000 | 0.932 | 0.936 | 0.001 | 0.937 | 0.980 | 0.000 | 0.928 | 0.915 |
| Moderate Correlation |  |  |  |  |  |  |  |  |  |  |  |  |  |
| 5 | BFL | 0.006 | 0.916 | 1.000 | 0.004 | 0.880 | 1.000 | 0.002 | 0.922 | 1.000 | 0.000 | 0.851 | 1.000 |
|  | BCL-OFS | 0.006 | 0.922 | 0.960 | 0.008 | 0.909 | 0.999 | 0.011 | 0.908 | 1.081 | 0.008 | 0.932 | 0.783 |
| 10 | BFL | 0.000 | 0.928 | 1.000 | 0.009 | 0.890 | 1.000 | 0.003 | 0.921 | 1.000 | 0.006 | 0.867 | 1.000 |
|  | BCL-OFS | 0.003 | 0.945 | 1.025 | 0.005 | 0.948 | 0.991 | 0.007 | 0.948 | 0.983 | 0.003 | 0.950 | 0.832 |
| 20 | BFL | 0.004 | 0.955 | 1.000 | 0.005 | 0.927 | 1.000 | 0.001 | 0.953 | 1.000 | 0.000 | 0.914 | 1.000 |
|  | BCL-OFS | 0.001 | 0.966 | 1.080 | 0.001 | 0.963 | 1.050 | 0.002 | 0.944 | 1.068 | 0.005 | 0.945 | 0.961 |
| 40 | BFL | 0.006 | 0.959 | 1.000 | 0.000 | 0.952 | 1.000 | 0.000 | 0.956 | 1.000 | 0.001 | 0.948 | 1.000 |
|  | BCL-OFS | 0.001 | 0.946 | 1.143 | 0.002 | 0.957 | 1.016 | 0.001 | 0.958 | 1.042 | 0.005 | 0.952 | 1.127 |
| 100 | BFL | 0.002 | 0.959 | 1.000 | 0.000 | 0.935 | 1.000 | 0.001 | 0.948 | 1.000 | 0.002 | 0.942 | 1.000 |
|  | BCL-OFS | 0.004 | 0.947 | 1.039 | 0.002 | 0.930 | 0.984 | 0.000 | 0.940 | 0.944 | 0.003 | 0.945 | 0.935 |
| Low Correlation |  |  |  |  |  |  |  |  |  |  |  |  |  |
| 5 | BFL | 0.009 | 0.938 | 1.000 | 0.013 | 0.879 | 1.000 | 0.001 | 0.927 | 1.000 | 0.007 | 0.894 | 1.000 |
|  | BCL-OFS | 0.018 | 0.929 | 1.017 | 0.016 | 0.933 | 0.913 | 0.009 | 0.927 | 0.984 | 0.010 | 0.939 | 0.836 |
| 10 | BFL | 0.005 | 0.943 | 1.000 | 0.012 | 0.905 | 1.000 | 0.010 | 0.935 | 1.000 | 0.005 | 0.908 | 1.000 |
|  | BCL-OFS | 0.009 | 0.947 | 1.071 | 0.003 | 0.949 | 1.000 | 0.005 | 0.968 | 0.976 | 0.004 | 0.958 | 0.872 |
| 20 | BFL | 0.007 | 0.947 | 1.000 | 0.003 | 0.935 | 1.000 | 0.004 | 0.961 | 1.000 | 0.002 | 0.927 | 1.000 |
|  | BCL-OFS | 0.002 | 0.962 | 1.011 | 0.001 | 0.957 | 1.029 | 0.003 | 0.959 | 1.051 | 0.010 | 0.948 | 0.957 |
| 40 | BFL | 0.008 | 0.956 | 1.000 | 0.000 | 0.942 | 1.000 | 0.002 | 0.968 | 1.000 | 0.002 | 0.953 | 1.000 |
|  | BCL-OFS | 0.002 | 0.957 | 1.027 | 0.001 | 0.964 | 0.944 | 0.002 | 0.965 | 0.920 | 0.001 | 0.961 | 0.925 |
| 100 | BFL | 0.005 | 0.954 | 1.000 | 0.001 | 0.944 | 1.000 | 0.000 | 0.952 | 1.000 | 0.003 | 0.949 | 1.000 |
|  | BCL-OFS | 0.001 | 0.940 | 0.993 | 0.002 | 0.946 | 0.938 | 0.001 | 0.931 | 1.066 | 0.000 | 0.941 | 0.938 |
| Varying Correlation |  |  |  |  |  |  |  |  |  |  |  |  |  |
| 5 | BFL | 0.003 | 0.917 | 1.000 | 0.009 | 0.869 | 1.000 | 0.027 | 0.927 | 1.000 | 0.012 | 0.862 | 1.000 |
|  | BCL-OFS | 0.005 | 0.926 | 0.945 | 0.007 | 0.915 | 0.969 | 0.010 | 0.936 | 1.004 | 0.006 | 0.924 | 0.856 |
| 10 | BFL | 0.000 | 0.932 | 1.000 | 0.016 | 0.897 | 1.000 | 0.001 | 0.918 | 1.000 | 0.001 | 0.911 | 1.000 |
|  | BCL-OFS | 0.006 | 0.940 | 1.068 | 0.001 | 0.943 | 0.931 | 0.013 | 0.961 | 0.966 | 0.010 | 0.943 | 1.018 |
| 20 | BFL | 0.005 | 0.930 | 1.000 | 0.007 | 0.917 | 1.000 | 0.001 | 0.940 | 1.000 | 0.002 | 0.930 | 1.000 |
|  | BCL-OFS | 0.006 | 0.958 | 0.978 | 0.002 | 0.960 | 0.972 | 0.001 | 0.964 | 1.056 | 0.006 | 0.947 | 1.025 |
| 40 | BFL | 0.003 | 0.951 | 1.000 | 0.004 | 0.936 | 1.000 | 0.006 | 0.959 | 1.000 | 0.001 | 0.937 | 1.000 |
|  | BCL-OFS | 0.002 | 0.953 | 1.084 | 0.005 | 0.958 | 0.890 | 0.003 | 0.964 | 1.071 | 0.002 | 0.958 | 1.066 |
| 100 | BFL | 0.004 | 0.951 | 1.000 | 0.003 | 0.941 | 1.000 | 0.001 | 0.961 | 1.000 | 0.002 | 0.952 | 1.000 |
|  | BCL-OFS | 0.000 | 0.928 | 1.022 | 0.001 | 0.929 | 0.980 | 0.000 | 0.934 | 1.114 | 0.003 | 0.933 | 1.025 |

Our exploration of computational efficiency is presented in Figure 2 and Table S7, which depicts the running time of various methods across different scenarios. The BCL method, with the parallel sampling strategy, consistently exhibited the fastest running time for all configurations. This finding highlighted the substantial performance improvement achieved by incorporating the parallel sampling strategy into the proposed BCL method. Conversely, the BFL methods, particularly those assuming unequal correlations, had the slowest running time. This can be attributed to the increasing number of parameters required by the BFL method to estimate variance-covariance structures as the number of treatments increased. Such a challenge was effectively addressed by the BCL method, which relied on simpler parameterizations, resulting in a more computationally efficient approach, especially for NMA datasets with a large number of treatments and studies.

**Figure 2.**
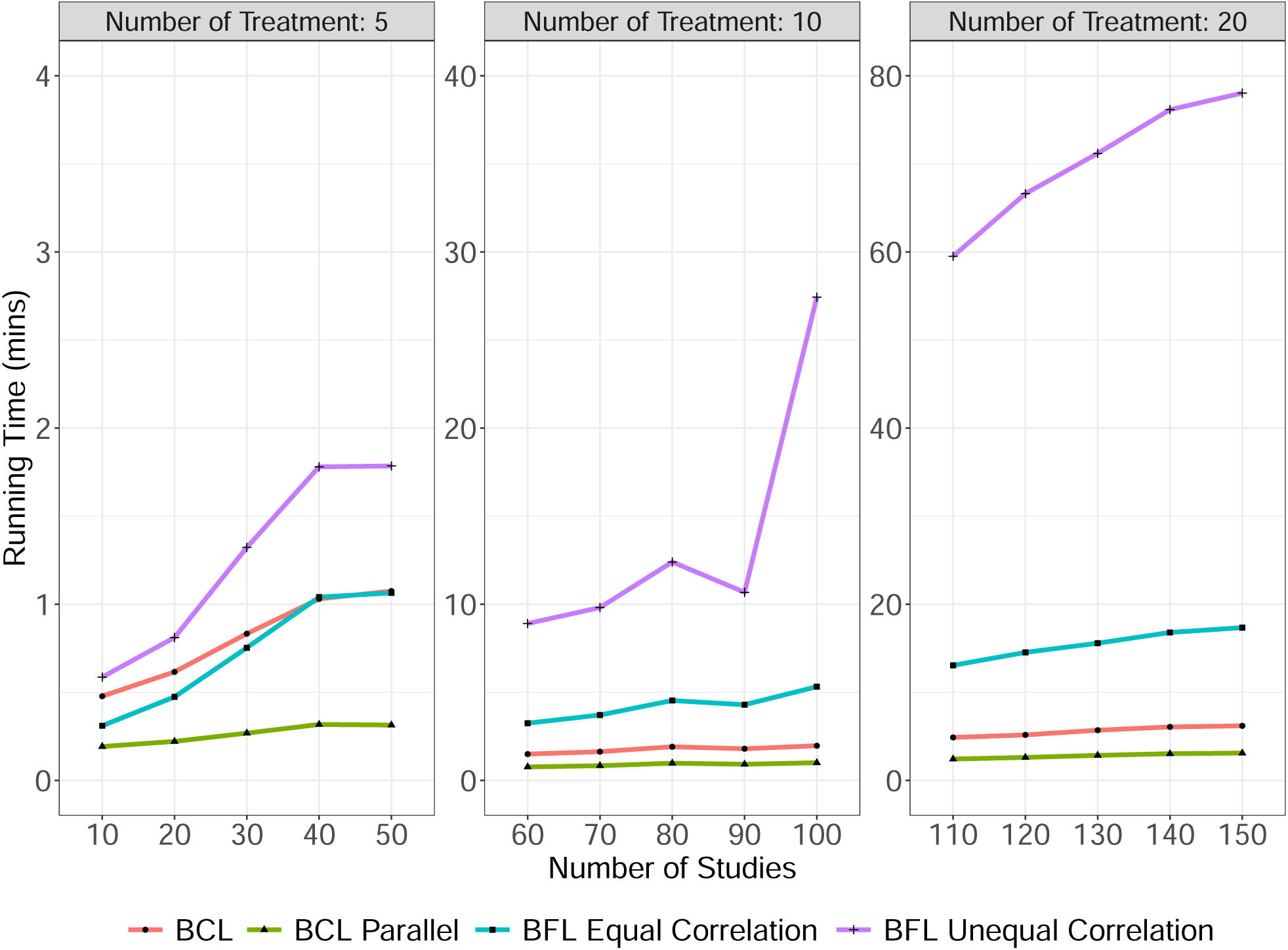
Comparisons of running time (in minutes) for the proposed method without and with the parallel sampling strategy (BCL/BCL Parallel), the Bayesian full-likelihood method with equal within-study correlations (BFL Equal Correlation), and the Bayesian full-likelihood method with unequal within-study correlations (BFL Unequal Correlation).

In summary, our simulation study demonstrated the unbiasedness and robustness of the proposed BCL method for multivariate NMAs. The BCL-OFS method achieved acceptable to good coverage probabilities. These findings indicated that the proposed method was robust under scenarios with both common and unequal between-study heterogeneity variances, as well as various within-study correlation structures. Moreover, the simulation study revealed significant computational efficiency gains with the proposed BCL method compared to existing BFL methods. Existing methods often require complex parameter specifications and suffer from substantially longer computational time, particularly for NMA datasets with increasing numbers of studies and treatments.

## 5. Applications to the two motivating examples

### 5.1 Application to root coverage

The MCMC setup and the potential scale reduction factor are provided in Appendices E and F of the Supplementary Materials. The NMA estimates and 90% CrIs for the four outcomes, CRC, RecRed, CAL gain, and KT gain, are displayed in Figures S6–S9 of the Supplementary Materials. For the outcomes of CRC, RecRed and KT gain, NMA results were consistent across both the BCL-OFS and the standard Bayesian NMA method. However, several discrepancies were observed for CAL gain. These discrepancies were exemplified by the results for CAF+CM versus CAF+EMD (BCL-OFS: MD = *−*0.26; 90% CrI, *−*1.20 to 0.68; the standard NMA: MD = 0.56; 90% CrI, *−*2.04 to 3.27), CAF versus CAF+EMD (BCL-OFS: MD = *−*0.21; 90% CrI, *−*0.73 to 0.31; the standard NMA: MD = 0.41; 90% CrI, *−*1.11 to 2.02), and CAF+CM versus CAF (BCL-OFS: MD = 0.06; 90% CrI, *−*0.50 to 0.60; the standard NMA: MD = *−*0.15; 90% CrI, *−*2.77 to 2.42). Moreover, the standard Bayesian method exhibited wider 90% CrIs. These discrepancies could likely be attributed to two reasons. First, the original analysis found network inconsistency, which contributed to the large variability in the results of both direct and indirect comparisons. Second, the BCL-OFS method was robust to misspecifed within-study correlations.

Figure 3 shows two-dimensional concordance plots of Z values, comparing the BCL and BCL-OFS methods to the standard NMA method across four relevant outcomes, where an absolute Z value of less than 1.96 corresponded to a p-value of less than 0.05. Most points fell within the diagonal quadrants, indicating agreement between the proposed and the standard NMA methods. Nevertheless, some points displayed discordant evidence of treatment comparisons for the outcome of CAL gain. We noted that some discrepancies appeared between the BCL and BCL-OFS methods. This difference might be attributed to the correction of standard errors for estimated pooled treatment effects in the BCL-OFS method.

**Figure 3.**
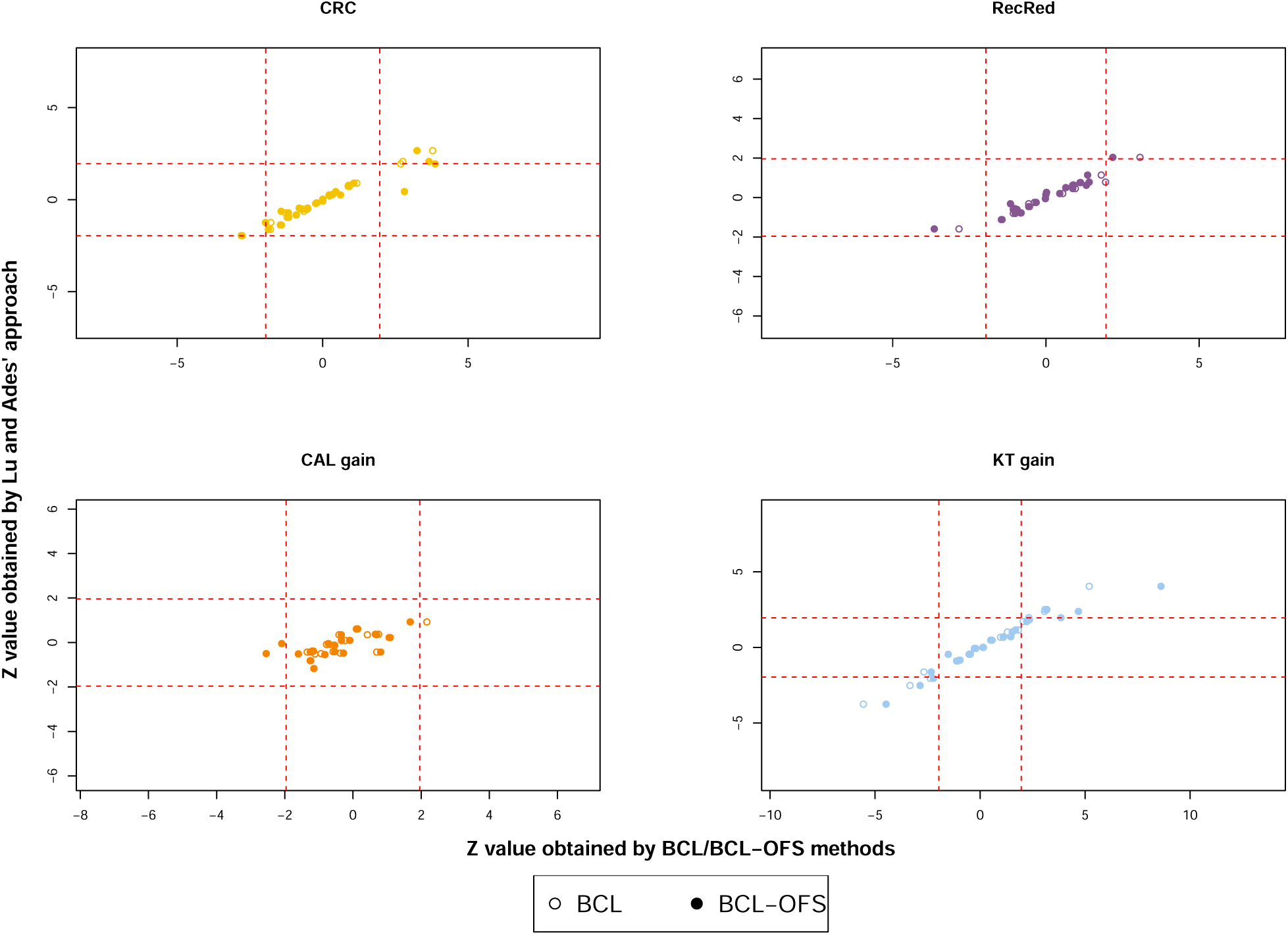
Concordance plots comparing Z values from the proposed BCL and BCL-OFS methods (x-axis) versus the standard Bayesian NMA using Lu and Ades’ method (y-axis) across four outcomes for the root coverage data. Open circles represent results from the BCL method versus the standard Bayesian NMA using Lu and Ades’ approach, while solid circles represent results from the BCL-OFS method versus the standard Bayesian NMA using Lu and Ades’ approach. Red dashed lines represent the critical values for 95% confidence levels.

Figure S5 of the Supplementary Materials presents the ranking of treatments obtained by integrating posterior estimates into the SUCRA calculation for four outcomes related to the root coverage data. As expected, the most effective treatment was CAF+CTG for RecRed (SUCRA = 0.87) and CAL gain (SUCRA = 0.94). For the outcome CRC, the most effective treatment was CAF+EMD (SUCRA = 0.99), while it was CAF+CM (SUCRA = 0.99) for KT gain. These results align with the findings of Buti et al. (2013). However, the BCL-OFS method revealed a key difference in the outcome CRC; the comparison between CAF+CTG and CAF+EMD was not statistically significant (OR = 0.61, 90% CrI, 0.24 to 1.52) in the standard NMA method but was significant (OR = 0.63, 90% CrI, 0.40 to 0.93) in the BCL-OFS method. This implies that CAF+CTG may not be the gold standard for the outcome CRC.

### 5.2 Application to anaemia in adults with chronic kidney disease

The league tables with NMA estimates and 95% CrIs (or CIs) for the two primary outcomes of interest, among a total of 9 outcomes, including preventing blood transfusion and allcause death, are summarized in Figures S12 and S13 of the Supplementary Materials. These estimates were obtained from the two methods: the standard NMA employing White’s approach (White et al., 2012), and the BCL-OFS method. Overall, both methods yielded largely consistent NMA estimates for the two primary outcomes. For the outcome of preventing blood transfusion, as shown in the upper triangular matrix of Figure S12, four ESAs demonstrated significant risk reduction in blood transfusion compared to placebo: darbepoetin alfa (OR = 0.37; 95% CrI, 0.13 to 0.82), Epoetin beta (OR = 0.25; 95% CrI, 0.10 to 0.52), methoxy polyethylene glycol-epoetin beta (OR = 0.35; 95% CrI, 0.10 to 0.91), and epoetin alfa (OR = 0.35; 95% CrI, 0.12 to 0.72). However, a discrepancy was observed between the two methods. When comparing methoxy polyethylene glycol-epoetin beta to placebo, the proposed method identified the significant result (OR = 0.35; 95% CrI, 0.10 to 0.91), while the White’s approach found the non-significant result (OR = 0.33; 95% CI, 0.11 to 1.02). For the outcome of all-cause death, NMA estimates revealed high consistency between the two methods, with only three exceptions in treatment comparisons, including methoxy polyethylene glycol-epoetin beta versus placebo, darbepoetin alfa versus methoxy polyethylene glycol-epoetin beta (Figure S13). These discrepancies may be attributed to the two reasons. First, the *I*^2^ statistic from the original analysis was 0% when comparing darbepoetin alfa to methoxy polyethylene glycol-epoetin beta. This indicated that the total variance was not completely dominated by the between-study variance, and some unexplained variability likely arose from within-study correlations. If these within-study correlations were not properly specified, they might result in overestimated standard errors of the pooled estimates for treatment comparisons. Second, methoxy polyethylene glycol-epoetin beta has very low certainty evidence from the original study, which might contribute to the observed inconsistencies.

Figure 4 displays two-dimensional concordance plots of statistical significance based on the Z values obtained from these methods, including the standard NMA using White’s method and the proposed BCL and BCL-OFS methods. These results revealed a high degree of concordance between the methods across nine outcomes. Most data points clustered around the diagonal quadrants, suggesting agreement in treatment comparison significance between the BCL method and the standard NMA method. However, some points from the proposed BCL-OFS method deviated from the diagonal for the outcome of major cardiovascular events.

**Figure 4.**
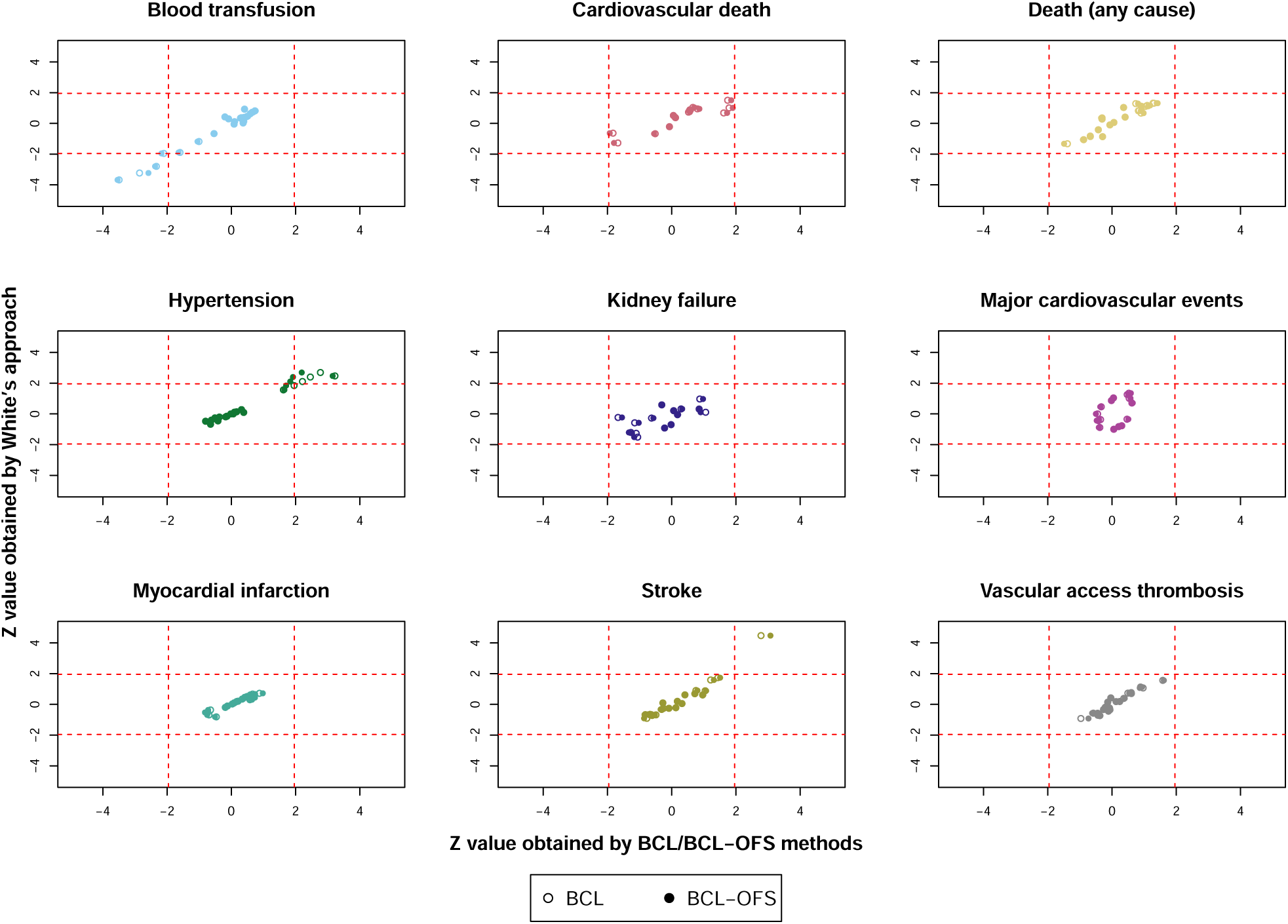
Concordance plots comparing Z values from the proposed BCL and BCL-OFS methods (x-axis) versus the standard NMA using White’s approach (y-axis) across all 9 outcomes for the anaemia data. Open circles represent results from the BCL method versus the standard NMA using White’s approach, while solid circles represent results from the BCL-OFS method versus the standard NMA using White’s approach. Red dashed lines represent the critical values for 95% confidence levels.

In Figure S11 of the Supplementary Materials, epoetin beta was selected as the topranked treatment based on SUCRA values, achieving the highest scores of 0.76 for preventing blood transfusion and 0.87 for all-cause death. These results were consistent with the previous research by Chung et al. (2023), indicating potential clinical benefits associated with epoetin beta. Interestingly, epoetin beta exhibited the lowest SUCRA value of 0.26 for the outcome of hypertension, aligning with earlier findings (Chung et al., 2023). Although the previous study did not definitively establish whether one epoetin drug was superior or safer than another, the highest SUCRA values of 0.99 among these outcomes suggested that epoetin beta held a distinct advantage in preventing cardiovascular death and myocardial infarction.

## 6. Discussion

This paper introduced a new method for analyzing multiple outcomes in NMAs. The proposed BCL method provided unbiased estimates of relative treatment effects without necessitating knowledge of within-study correlations among treatments and outcomes. An efficient hybrid Gibbs sampler was implemented to draw samples from the posterior distributions, which captured the uncertainty around the estimated treatment effects. Furthermore, we introduced the OFS method applied to these posterior samples to provide a better approximation of the true distributions of treatment effects. Extensive simulations demonstrated the robustness of the BCL and BCL-OFS methods under various scenarios, including the number of studies, within-study and between-study correlations, and both homogeneous and heterogeneous between-study variances. Based on the simulation results, we recommended applying the proposed BCL-OFS method for scenarios with at least low to moderate and higher, as well as varying correlation settings.

The proposed method offers several advantages. First, it eliminates the need to specify within-study correlations, leading to valid statistical inferences regardless of the underlying correlation structures, as demonstrated in our simulation study. Second, it estimates both treatment effects and between-study variance only, leading to a more parsimonious parameter set compared to the Bayesian full-likelihood methods. This, coupled with the parallel sampling strategy, translates to computational efficiency for large and complex NMAs with an increasing number of studies and treatments. Third, while incorporating multiple outcomes introduces complications in ensuring a positive semi-definite constraint on the variance-covariance matrix (Nam et al., 2003), our proposed method can mitigate such constraints. Finally, employing the OFS adjustment aids in calibrating the posterior samples and approximating the true posterior distributions. In contrast to existing in-sampling location and curvature adjustment methods (Pauli et al., 2011), the OFS adjustment reduces computational cost while avoiding potential distortions of the MCMCs during the sampling process.

While the proposed method demonstrates numerous advantages, it also offers opportunities for further exploration in several areas. First, as network complexity grows, the computational cost associated with the OFS adjustment, particularly for calculating the score vector and Hessian matrix, can become substantial. This motivates the investigation of computationally efficient extensions to the OFS method, thereby accommodating its applicability in large-scale network meta-analyses. Second, the current implementation of the Gibbs sampler employs a uniform step size across all outcomes. Incorporating an adaptive MH algorithm (Brooks et al., 2011) could significantly improve efficiency by allowing self-adaptive step sizes for each outcome, leading to faster convergence and more accurate estimates. Finally, the motivating example highlighted large variability, potentially attributable to unreported outcomes in the network datasets. In practice, outcome reporting bias, where selective reporting of outcomes can occur, is a recognized concern (Marks-Anglin and Chen, 2020). The proposed method may be extended to incorporate a sensitivity analysis (Hutton and Williamson, 2000) to account for this potential bias.

## Data Availability

All data used in this work are available online at
DOI: 10.1111/jcpe.12028
DOI: 10.1002/14651858.CD010590.pub3

https://doi.org/10.1111/jcpe.12028

https://doi.org/10.1002/14651858.CD010590.pub3

## Acknowledgements

This work was supported by NIH grant: R01DK128237 (YL and YW).

## Supporting Information

The supplementary materials are available under the Paper Information link at the *Biometrics* website http:www.tibs.org/biometrics.

