## Supplementary material for "Exploiting multivariate network meta-analysis: A calibrated Bayesian composite likelihood inference": NA

### Web-based Supplementary Materials for "Exploiting multivariate network meta-analysis: A calibrated Bayesian composite likelihood inference" by

Yifei Wang, Lifeng Lin, and Yu-Lun Liu

**Appendix A: Networks of evidence diagrams**

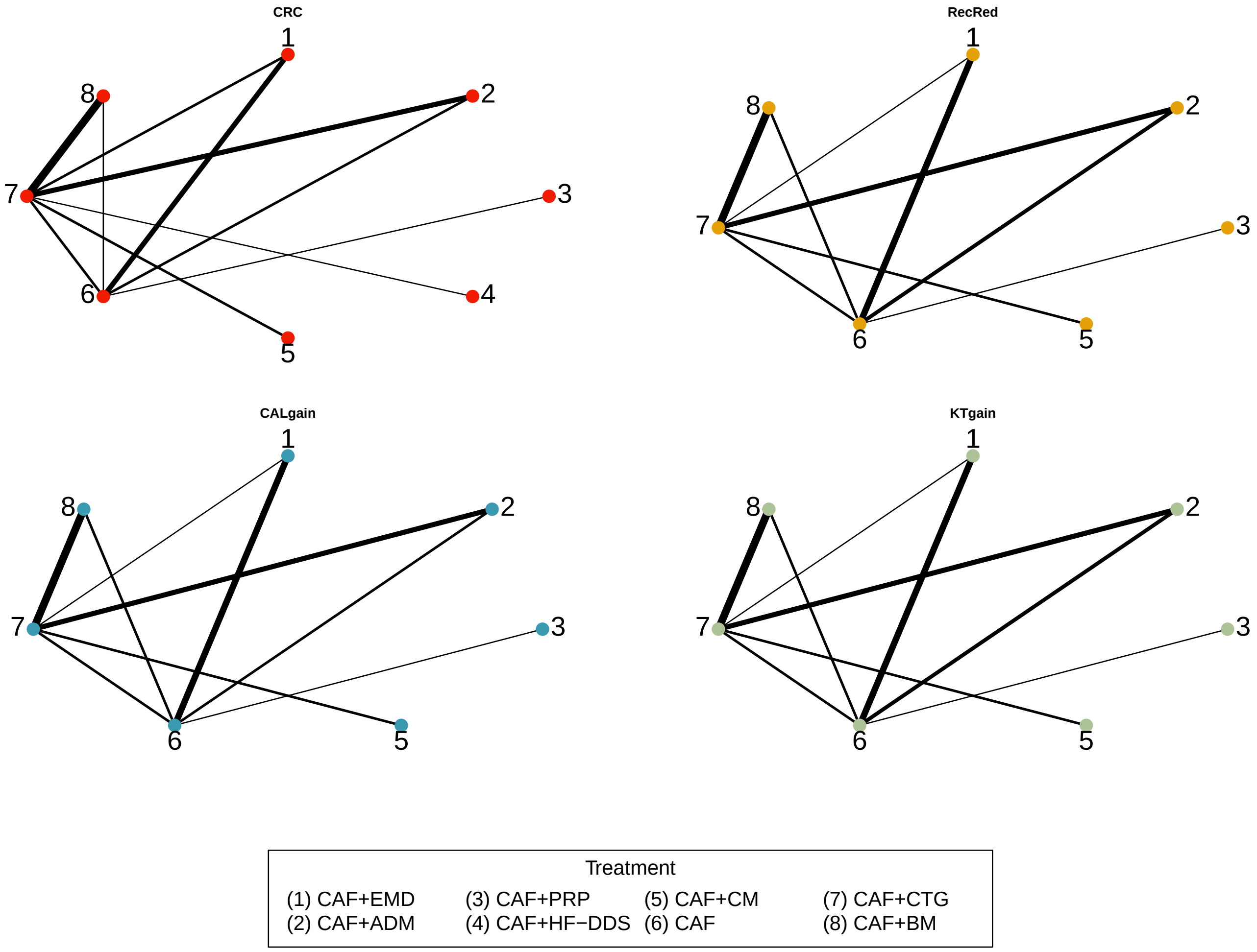


Web Figure S1: Illustration of evidence network diagrams for root coverage treatments. The size of the node is proportional to the number of participants assigned to each treatment. Solid lines represent direct comparisons with trials, and the thickness of a line connecting nodes is proportional to the number of trials directly comparing each pair of treatments.

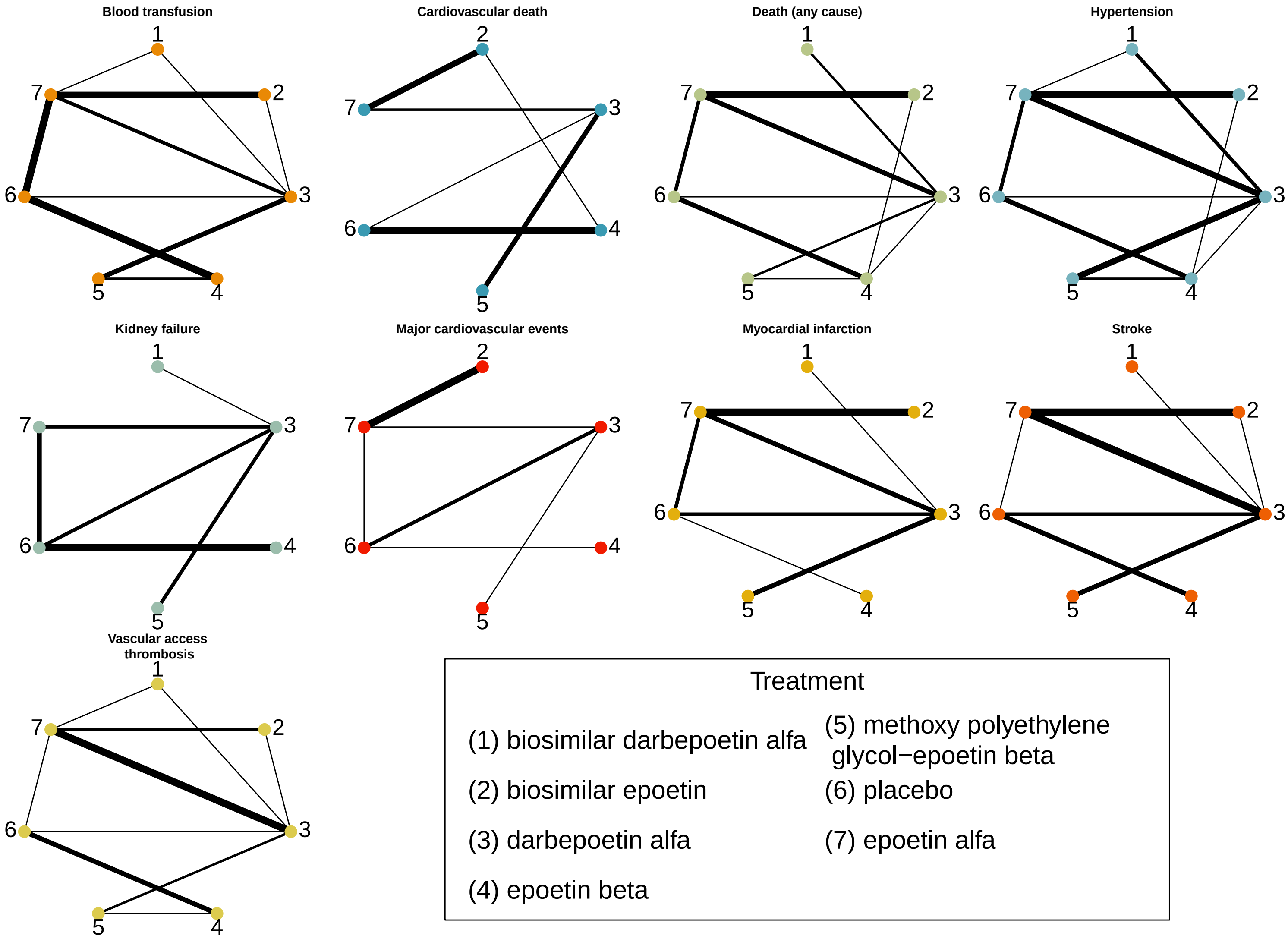


Web Figure S2: Illustration of evidence network diagrams for ESA treatments. The size of the node is proportional to the number of participants assigned to each treatment. Solid lines represent direct comparisons with trials, and the thickness of a line connecting nodes is proportional to the number of trials directly comparing each pair of treatments.

#### Appendix B: Derivation of posterior distributions for treatment effect parameters

Given the treatment effect $\mu_l^{XZ}$ for a comparison between treatments $X$ and $Z$ on $l$ outcome, the observed data $\mathbf{y}$, the within-study variance $s_{i,l}^{XZ^2}$ and the between-study variance $\tau_l^{XZ^2}$, the composite likelihood function is

$$L\left(\tau_l^{XZ^2}, \mu_l^{XZ}\right) \propto \left\{\prod_{i \in \mathcal{A}_l^{XZ}} \left(s_{i,l}^{XZ^2} + \tau_l^{XZ^2}\right)^{-1/2}\right\} \times \exp\left\{\sum_{i \in \mathcal{A}_l^{XZ}} -\frac{\left(y_{i,l}^{XZ} - \mu_l^{XZ}\right)^2}{2\left(s_{i,l}^{XZ^2} + \tau_l^{XZ^2}\right)}\right\},$$

where the set $A_l^{XZ}$ includes the studies comparing treatment $X$ and treatment $Z$ with respect to outcome $l$. The posterior kernel function is given by

$$\begin{aligned}
P\left(\mu_l^{XZ} \mid \tau, \mathbf{y}\right) &\propto P\left(\mathbf{y} \mid \tau_l^{XZ^2}, \mu_l^{XZ}\right) P\left(\mu_l^{XZ}\right) \propto L\left(\tau_l^{XZ^2}, \mu_l^{XZ}\right) P\left(\mu_l^{XZ}\right) \\
&\propto \exp\left\{\sum_{i \in \mathcal{A}_l^{XZ}} -\frac{\left(y_{i,l}^{XZ} - \mu_l^{XZ}\right)^2}{2\left(s_{i,l}^{XZ^2} + \tau_l^{XZ^2}\right)}\right\} \times \exp\left\{-\frac{\left(\mu_l^{XZ}\right)^2}{20000}\right\} \\
&\propto \exp\left\{\sum_{i \in A_l^{XZ}} -\frac{-2y_{i,l}^{X}\mu_l^{XZ} + \mu_l^{XZ^2}}{2\left(s_{i,l}^{XZ^2} + \tau_l^{XZ^2}\right)}\right\} \times \exp\left(-\frac{\left(\mu_l^{XZ}\right)^2}{20000}\right) \\
&\propto \exp\left[-\left\{\frac{1}{20000} + \sum_{i \in A_l^{XZ}} \frac{1}{2\left(s_{i,l}^{XZ^2} + \tau_l^{XZ^2}\right)}\right\} \mu_l^{XZ^2} + \right. \\
&\qquad \left. \left\{\sum_{i \in A_l^{XZ}} \frac{y_{i,l}^{XZ}}{\left(s_{i,l}^{XZ^2} + \tau_l^{XZ^2}\right)}\right\} \mu_l^{XZ}\right] \\
&\propto \exp\left(-A\mu_l^{XZ^2} + B\mu_l^{XZ}\right) \\
&\propto \exp\left(-\frac{\mu_l^{XZ^2} - 2\frac{B}{2A}\mu_l^{XZ}}{1/A}\right) \\
&\propto \exp\left\{-\frac{\left(\mu_l^{XZ} - \frac{B}{2A}\right)^2}{1/A}\right\}
\end{aligned}$$

Therefore, the posterior kernel function of $\mu_l^{XZ}$ aligns with a normal distribution $\mathrm{N}\left(\frac{B}{2A}, \frac{1}{2A}\right)$, where

$$A = \frac{1}{20000} + \sum_{i \in A_l^{XZ}} \frac{1}{2\left(s_{i,l}^{XZ^2} + \tau_l^{XZ^2}\right)};$$

$$B = \sum_{i \in A_l^{XZ}} \frac{y_{i,l}^{XZ}}{\left(s_{i,l}^{XZ^2} + \tau_l^{XZ^2}\right)}.$$

#### Appendix C: Simulation design

We assumed a network with three treatments, denoted $A$, $B$, and $C$, where $A$ is treated as the reference treatment. We set up two continuous outcomes $l \in \{1, 2\}$, and three relative treatment effects: $AB$, $BC$, and $AC$. In the simulation study, data for each study $i$ were generated from a multivariate normal distribution:

$$\mathbf{y}_i = (y_{i,1}^{AB}, y_{i,1}^{AC}, y_{i,2}^{AB}, y_{i,2}^{AC})^\top \sim \left( \left(\mu_1^{AB}, \mu_1^{AC}, \mu_2^{AB}, \mu_2^{AC}\right)^\top, \mathbf{D}^b\mathbf{R}^b\mathbf{D}^b + \mathbf{D}^w\mathbf{R}^w\mathbf{D}^w \right);$$
$$\mathbf{D}^b = \text{diag}\left( \left[ \tau_1^{AB^2}, \tau_1^{AC^2}, \tau_2^{AB^2}, \tau_2^{AC^2} \right] \right);$$
$$\mathbf{D}^w = \text{diag}\left( \left[ s_{i,1}^{AB^2}, s_{i,1}^{AC^2}, s_{i,2}^{AB^2}, s_{i,2}^{AC^2} \right] \right).$$

The between-study correlation matrix $\mathbf{R}^b$ and the within-study correlation matrix $\mathbf{R}^w$ are

$$\mathbf{R}^b = \begin{pmatrix} 1 & \rho^b & \rho^b & \rho^b \\ \rho^b & 1 & \rho^b & \rho^b \\ \rho^b & \rho^b & 1 & \rho^b \\ \rho^b & \rho^b & \rho^b & 1 \end{pmatrix} \text{ and } \mathbf{R}^w = \begin{pmatrix} 1 & \rho^w & \rho^w & \rho^w \\ \rho^w & 1 & \rho^w & \rho^w \\ \rho^w & \rho^w & 1 & \rho^w \\ \rho^w & \rho^w & \rho^w & 1 \end{pmatrix}.$$

By assuming consistency, the relative treatment effect $\mu_l^{BC}$ is calculated from $\mu_l^{AB}$ and $\mu_l^{AC}$, and the between-study variance $\tau_l^{BC^2}$ is calculated from $\tau_l^{AB^2}$ and $\tau_l^{AC^2}$ for $l \in \{1, 2\}$

$$\mu_l^{BC} = \mu_l^{AB} - \mu_l^{AC};$$
$$\tau_l^{BC} = \tau_l^{AB^2} + \tau_l^{AC^2} - 2\rho^b\tau_l^{AB}\tau_l^{AC}.$$

In a common between-study variance scenario, we set $\tau_l^{XZ^2} = 0.36$ for the outcome $l \in \{1, 2\}$ and the treatment effect $XZ \in \{AB, BC, AC\}$. In this scenario, we considered the number of studies, including 5, 10, 20, 40, and 100. Moreover, we examined the impact of different correlation structures: high correlations ($\rho^w = \rho^b = 0.8$), moderate correlations ($\rho^w = \rho^b = 0.5$), low correlations ($\rho^w = \rho^b = 0.2$), and varying correlations. In a varying correlation setting, correlations were set as follows:

$$\mathbf{R}^b = \mathbf{R}^w = \begin{bmatrix} 1.0 & 0.5 & 0.4 & 0.3 \\ 0.5 & 1.0 & 0.5 & 0.4 \\ 0.4 & 0.5 & 1.0 & 0.3 \\ 0.3 & 0.4 & 0.3 & 1.0 \end{bmatrix}.$$

Other parameter settings are

$$(\mu_1^{AB}, \mu_1^{AC}, \mu_2^{AB}, \mu_2^{AC})^\top = (1, 2, -1, -2)^\top;$$
$$s_{i,1}^{AB^2} = s_{i,1}^{AC^2} = s_{i,1}^{BC^2} = s_{i,2}^{AB^2} = s_{i,2}^{AC^2} = s_{i,2}^{BC^2} = 1.$$

In the unequal between-study variance scenario, the parameters were specified as follows:

$$(\tau_1^{AB^2}, \tau_1^{AC^2}, \tau_2^{AB^2}, \tau_2^{AC^2})^\top = (0.25, 0.36, 0.36, 0.49)^\top.$$

The number of studies and other parameter settings were the same as in the previous scenario. For each scenario, we generated $1,000$ datasets and applied both the proposed methods, BCL and BCL-OFS. We assessed the performance of the proposed methods by examining bias and coverage probability.

#### Appendix D: Additional results for simulation studies

Web Table S1: Summary of $1,000$ simulations with $I = 5$, 10, 20, 40 and 100: bias and coverage probability (CP) of pooled estimates of $AB$ and $AC$ treatment comparisons for two outcomes. The data generating mechanism was through an unequal between-study heterogeneity variance with ${\tau_1^{AB}}^2 = 0.25$, ${\tau_1^{AC}}^2 = 0.36$, ${\tau_2^{AB}}^2 = 0.36$, ${\tau_2^{AC}}^2 = 0.49$, $\mu_1^{AB} = 1$, $\mu_1^{AC} = 2$, $\mu_2^{AB} = -1$, and $\mu_2^{AC} = -2$, as well as high correlations $\rho^w = \rho^b = 0.8$, moderate correlations $\rho^w = \rho^b = 0.5$, low correlations $\rho^w = \rho^b = 0.2$, and varying correlations. Results were based on the proposed BCL method.

| | High correlation | | | | Moderate correlation | | | | Low correlation | | | | Varying correlation | | | |
|---|---|---|---|---|---|---|---|---|---|---|---|---|---|---|---|---|
| | $\mu_1^{AB}$ | $\mu_1^{AC}$ | $\mu_2^{AB}$ | $\mu_2^{AC}$ | $\mu_1^{AB}$ | $\mu_1^{AC}$ | $\mu_2^{AB}$ | $\mu_2^{AC}$ | $\mu_1^{AB}$ | $\mu_1^{AC}$ | $\mu_2^{AB}$ | $\mu_2^{AC}$ | $\mu_1^{AB}$ | $\mu_1^{AC}$ | $\mu_2^{AB}$ | $\mu_2^{AC}$ |
| **I=5** | | | | | | | | | | | | | | | | |
| Bias | 0.004 | 0.000 | 0.003 | 0.004 | 0.016 | 0.007 | 0.006 | 0.005 | 0.004 | 0.014 | 0.000 | 0.004 | 0.003 | 0.009 | 0.022 | 0.008 |
| CP | 0.917 | 0.907 | 0.892 | 0.882 | 0.922 | 0.922 | 0.914 | 0.893 | 0.912 | 0.930 | 0.910 | 0.889 | 0.931 | 0.917 | 0.926 | 0.916 |
| **I=10** | | | | | | | | | | | | | | | | |
| Bias | 0.001 | 0.003 | 0.001 | 0.004 | 0.001 | 0.006 | 0.002 | 0.005 | 0.010 | 0.006 | 0.005 | 0.009 | 0.007 | 0.010 | 0.011 | 0.001 |
| CP | 0.900 | 0.895 | 0.879 | 0.876 | 0.915 | 0.895 | 0.903 | 0.892 | 0.920 | 0.916 | 0.942 | 0.910 | 0.895 | 0.905 | 0.894 | 0.898 |
| **I=20** | | | | | | | | | | | | | | | | |
| Bias | 0.006 | 0.004 | 0.005 | 0.004 | 0.004 | 0.009 | 0.004 | 0.007 | 0.003 | 0.013 | 0.006 | 0.004 | 0.004 | 0.005 | 0.002 | 0.001 |
| CP | 0.890 | 0.885 | 0.893 | 0.880 | 0.897 | 0.893 | 0.905 | 0.892 | 0.921 | 0.905 | 0.890 | 0.903 | 0.890 | 0.907 | 0.906 | 0.903 |
| **I=40** | | | | | | | | | | | | | | | | |
| Bias | 0.002 | 0.003 | 0.003 | 0.001 | 0.005 | 0.000 | 0.008 | 0.003 | 0.003 | 0.005 | 0.002 | 0.004 | 0.005 | 0.005 | 0.003 | 0.009 |
| CP | 0.895 | 0.873 | 0.885 | 0.892 | 0.901 | 0.892 | 0.910 | 0.893 | 0.896 | 0.886 | 0.914 | 0.900 | 0.900 | 0.899 | 0.896 | 0.923 |
| **I=100** | | | | | | | | | | | | | | | | |
| Bias | 0.001 | 0.002 | 0.001 | 0.003 | 0.001 | 0.002 | 0.000 | 0.001 | 0.002 | 0.002 | 0.000 | 0.000 | 0.002 | 0.002 | 0.001 | 0.000 |
| CP | 0.899 | 0.882 | 0.906 | 0.899 | 0.893 | 0.884 | 0.898 | 0.902 | 0.921 | 0.901 | 0.918 | 0.912 | 0.900 | 0.906 | 0.920 | 0.909 |

Web Table S2: Summary of $1,000$ simulations with $I = 5$, 10, 20, 40 and 100: bias and coverage probability (CP) of pooled estimates of $AB$ and $AC$ treatment comparisons for two outcomes. The data generating mechanism was through an unequal between-study heterogeneity variance with ${\tau_1^{AB}}^2 = 0.25$, ${\tau_1^{AC}}^2 = 0.36$, ${\tau_2^{AB}}^2 = 0.36$, ${\tau_2^{AC}}^2 = 0.49$, $\mu_1^{AB} = 1$, $\mu_1^{AC} = 2$, $\mu_2^{AB} = -1$, and $\mu_2^{AC} = -2$, as well as high correlations $\rho^w = \rho^b = 0.8$, moderate correlations $\rho^w = \rho^b = 0.5$, low correlations $\rho^w = \rho^b = 0.2$, and varying correlations. Results were based on the proposed BCL-OFS method.

| | High correlation | | | | Moderate correlation | | | | Low correlation | | | | Varying correlation | | | |
|---|---|---|---|---|---|---|---|---|---|---|---|---|---|---|---|---|
| | $\mu_1^{AB}$ | $\mu_1^{AC}$ | $\mu_2^{AB}$ | $\mu_2^{AC}$ | $\mu_1^{AB}$ | $\mu_1^{AC}$ | $\mu_2^{AB}$ | $\mu_2^{AC}$ | $\mu_1^{AB}$ | $\mu_1^{AC}$ | $\mu_2^{AB}$ | $\mu_2^{AC}$ | $\mu_1^{AB}$ | $\mu_1^{AC}$ | $\mu_2^{AB}$ | $\mu_2^{AC}$ |
| **I=5** | | | | | | | | | | | | | | | | |
| Bias | 0.004 | 0.000 | 0.003 | 0.004 | 0.016 | 0.007 | 0.006 | 0.005 | 0.004 | 0.014 | 0.000 | 0.004 | 0.003 | 0.009 | 0.022 | 0.008 |
| CP | 0.917 | 0.927 | 0.918 | 0.925 | 0.934 | 0.934 | 0.931 | 0.918 | 0.927 | 0.925 | 0.926 | 0.919 | 0.941 | 0.927 | 0.929 | 0.927 |
| **I=10** | | | | | | | | | | | | | | | | |
| Bias | 0.001 | 0.003 | 0.001 | 0.004 | 0.001 | 0.006 | 0.002 | 0.005 | 0.010 | 0.006 | 0.005 | 0.009 | 0.007 | 0.010 | 0.011 | 0.001 |
| CP | 0.946 | 0.941 | 0.935 | 0.941 | 0.956 | 0.937 | 0.952 | 0.954 | 0.945 | 0.944 | 0.973 | 0.958 | 0.941 | 0.961 | 0.947 | 0.948 |
| **I=20** | | | | | | | | | | | | | | | | |
| Bias | 0.006 | 0.004 | 0.005 | 0.004 | 0.004 | 0.009 | 0.004 | 0.007 | 0.003 | 0.013 | 0.006 | 0.004 | 0.004 | 0.005 | 0.002 | 0.001 |
| CP | 0.940 | 0.941 | 0.945 | 0.945 | 0.953 | 0.951 | 0.956 | 0.963 | 0.957 | 0.969 | 0.961 | 0.953 | 0.957 | 0.961 | 0.953 | 0.960 |
| **I=40** | | | | | | | | | | | | | | | | |
| Bias | 0.002 | 0.003 | 0.003 | 0.001 | 0.005 | 0.000 | 0.008 | 0.003 | 0.003 | 0.005 | 0.002 | 0.004 | 0.005 | 0.005 | 0.003 | 0.009 |
| CP | 0.956 | 0.947 | 0.936 | 0.934 | 0.954 | 0.944 | 0.956 | 0.947 | 0.950 | 0.949 | 0.964 | 0.950 | 0.952 | 0.955 | 0.947 | 0.964 |
| **I=100** | | | | | | | | | | | | | | | | |
| Bias | 0.001 | 0.002 | 0.001 | 0.003 | 0.001 | 0.002 | 0.000 | 0.001 | 0.002 | 0.002 | 0.000 | 0.000 | 0.002 | 0.002 | 0.001 | 0.000 |
| CP | 0.945 | 0.914 | 0.924 | 0.921 | 0.944 | 0.934 | 0.911 | 0.924 | 0.957 | 0.942 | 0.940 | 0.937 | 0.949 | 0.941 | 0.931 | 0.920 |

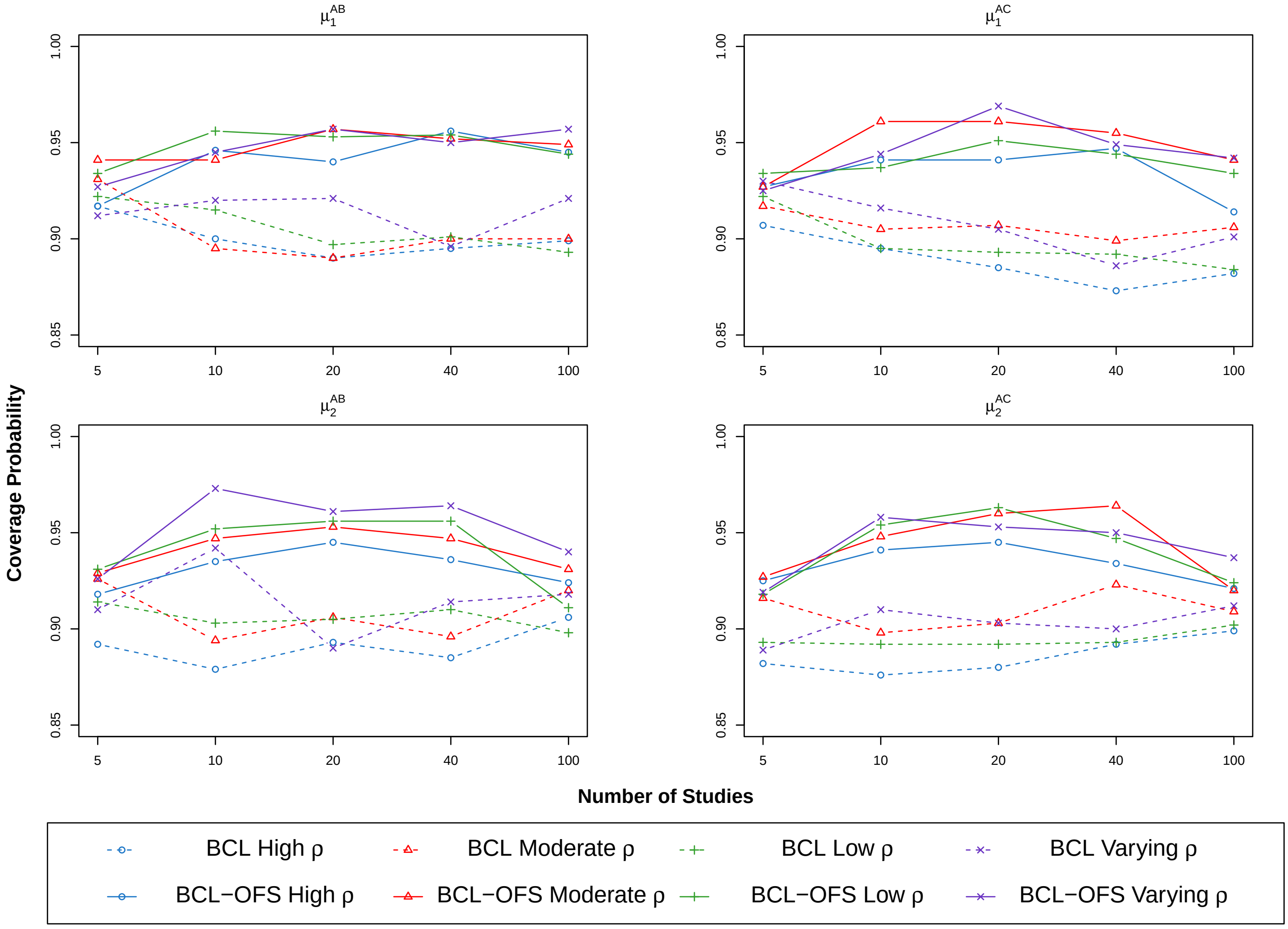


Web Figure S3: Coverage probabilities for pooled estimates of $AB$ and $AC$ treatment comparisons for two outcomes using the proposed BCL and BCL-OFS methods under low ($\rho^w = \rho^b = 0.2$), moderate ($\rho^w = \rho^b = 0.5$), high ($\rho^w = \rho^b = 0.8$), and varying correlations. The data generating mechanism was through an unequal between-study heterogeneity variance with ${\tau_1^{AB}}^2 = 0.25$, ${\tau_1^{AC}}^2 = 0.36$, ${\tau_2^{AB}}^2 = 0.36$, and ${\tau_2^{AC}}^2 = 0.49$.

Web Table S3: Summary of $1,000$ simulations with $I = 5$, 10, 20, 40 and 100: bias, coverage probability (CP) and relative efficiency (RE) of pooled estimates of $AB$ and $AC$ treatment comparisons for two outcomes. The data generating mechanism was through an unequal between-study heterogeneity variance with ${\tau_1^{AB}}^2 = 0.25$, ${\tau_1^{AC}}^2 = 0.36$, ${\tau_2^{AB}}^2 = 0.36$, ${\tau_2^{AC}}^2 = 0.49$, $\mu_1^{AB} = 1$, $\mu_1^{AC} = 2$, $\mu_2^{AB} = -1$, and $\mu_2^{AC} = -2$, as well as high correlations $\rho^w = \rho^b = 0.8$. Results were based on the BFL method and the proposed BCL-OFS method.)

| | | $\mu_1^{AB}$ | | | $\mu_1^{AC}$ | | | $\mu_2^{AB}$ | | | $\mu_2^{AC}$ | | |
|---|---|---|---|---|---|---|---|---|---|---|---|---|---|
| # Study | Method | Bias | CP | RE | Bias | CP | RE | Bias | CP | RE | Bias | CP | RE |
| 5 | BFL | 0.001 | 0.928 | 1.000 | 0.004 | 0.854 | 1.000 | 0.008 | 0.907 | 1.000 | 0.015 | 0.836 | 1.000 |
| | BCL-OFS | 0.004 | 0.917 | 1.038 | 0.000 | 0.927 | 0.883 | 0.003 | 0.918 | 1.099 | 0.004 | 0.925 | 0.919 |
| 10 | BFL | 0.007 | 0.923 | 1.000 | 0.008 | 0.873 | 1.000 | 0.004 | 0.929 | 1.000 | 0.019 | 0.870 | 1.000 |
| | BCL-OFS | 0.001 | 0.946 | 1.126 | 0.003 | 0.941 | 0.985 | 0.001 | 0.935 | 1.189 | 0.004 | 0.941 | 1.003 |
| 20 | BFL | 0.000 | 0.955 | 1.000 | 0.000 | 0.923 | 1.000 | 0.003 | 0.940 | 1.000 | 0.011 | 0.918 | 1.000 |
| | BCL-OFS | 0.006 | 0.940 | 1.091 | 0.004 | 0.941 | 1.022 | 0.005 | 0.945 | 1.045 | 0.004 | 0.945 | 1.069 |
| 40 | BFL | 0.005 | 0.956 | 1.000 | 0.002 | 0.927 | 1.000 | 0.001 | 0.961 | 1.000 | 0.003 | 0.932 | 1.000 |
| | BCL-OFS | 0.002 | 0.956 | 1.079 | 0.003 | 0.947 | 1.052 | 0.003 | 0.936 | 1.086 | 0.001 | 0.934 | 1.018 |
| 100 | BFL | 0.002 | 0.968 | 1.000 | 0.001 | 0.946 | 1.000 | 0.001 | 0.960 | 1.000 | 0.000 | 0.943 | 1.000 |
| | BCL-OFS | 0.001 | 0.945 | 1.070 | 0.002 | 0.914 | 1.102 | 0.001 | 0.924 | 1.048 | 0.003 | 0.921 | 1.049 |

Web Table S4: Summary of $1,000$ simulations with $I = 5$, 10, 20, 40 and 100: bias, coverage probability (CP) and relative efficiency (RE) of pooled estimates of $AB$ and $AC$ treatment comparisons for two outcomes. The data generating mechanism was through an unequal between-study heterogeneity variance with ${\tau_1^{AB}}^2 = 0.25$, ${\tau_1^{AC}}^2 = 0.36$, ${\tau_2^{AB}}^2 = 0.36$, ${\tau_2^{AC}}^2 = 0.49$, $\mu_1^{AB} = 1$, $\mu_1^{AC} = 2$, $\mu_2^{AB} = -1$, and $\mu_2^{AC} = -2$, as well as moderate correlations $\rho^w = \rho^b = 0.5$. Results were based on the BFL method and the proposed BCL-OFS method.

| | | $\mu_1^{AB}$ | | | $\mu_1^{AC}$ | | | $\mu_2^{AB}$ | | | $\mu_2^{AC}$ | | |
|---|---|---|---|---|---|---|---|---|---|---|---|---|---|
| # Study | Method | Bias | CP | RE | Bias | CP | RE | Bias | CP | RE | Bias | CP | RE |
| 5 | BFL | 0.003 | 0.916 | 1.000 | 0.019 | 0.862 | 1.000 | 0.016 | 0.918 | 1.000 | 0.007 | 0.863 | 1.000 |
| | BCL-OFS | 0.016 | 0.934 | 0.893 | 0.007 | 0.934 | 0.839 | 0.006 | 0.931 | 0.942 | 0.005 | 0.918 | 0.951 |
| 10 | BFL | 0.005 | 0.933 | 1.000 | 0.011 | 0.896 | 1.000 | 0.012 | 0.921 | 1.000 | 0.011 | 0.888 | 1.000 |
| | BCL-OFS | 0.001 | 0.956 | 0.984 | 0.006 | 0.937 | 1.033 | 0.002 | 0.952 | 0.873 | 0.005 | 0.954 | 0.919 |
| 20 | BFL | 0.006 | 0.955 | 1.000 | 0.004 | 0.929 | 1.000 | 0.007 | 0.953 | 1.000 | 0.008 | 0.943 | 1.000 |
| | BCL-OFS | 0.004 | 0.953 | 1.128 | 0.009 | 0.951 | 0.972 | 0.004 | 0.956 | 1.120 | 0.007 | 0.963 | 1.046 |
| 40 | BFL | 0.008 | 0.956 | 1.000 | 0.005 | 0.938 | 1.000 | 0.005 | 0.958 | 1.000 | 0.003 | 0.931 | 1.000 |
| | BCL-OFS | 0.005 | 0.954 | 1.064 | 0.000 | 0.944 | 1.065 | 0.008 | 0.956 | 1.040 | 0.003 | 0.947 | 1.141 |
| 100 | BFL | 0.005 | 0.959 | 1.000 | 0.000 | 0.943 | 1.000 | 0.000 | 0.963 | 1.000 | 0.004 | 0.940 | 1.000 |
| | BCL-OFS | 0.001 | 0.944 | 1.092 | 0.002 | 0.934 | 1.097 | 0.000 | 0.911 | 1.105 | 0.001 | 0.924 | 1.069 |

Web Table S5: Summary of 1,000 simulations with $I = 5$, 10, 20, 40 and 100: bias, coverage probability (CP) and relative efficiency (RE) of pooled estimates of $AB$ and $AC$ treatment comparisons for two outcomes. The data generating mechanism was through an unequal between-study heterogeneity variance with ${\tau_1^{AB}}^2 = 0.25$, ${\tau_1^{AC}}^2 = 0.36$, ${\tau_2^{AB}}^2 = 0.36$, ${\tau_2^{AC}}^2 = 0.49$, $\mu_1^{AB} = 1$, $\mu_1^{AC} = 2$, $\mu_2^{AB} = -1$, and $\mu_2^{AC} = -2$, as well as low correlations $\rho^w = \rho^b = 0.2$. Results were based on the BFL method and the proposed BCL-OFS method.

| | | $\mu_1^{AB}$ | | | $\mu_1^{AC}$ | | | $\mu_2^{AB}$ | | | $\mu_2^{AC}$ | | |
|---|---|---|---|---|---|---|---|---|---|---|---|---|---|
| # Study | Method | Bias | CP | RE | Bias | CP | RE | Bias | CP | RE | Bias | CP | RE |
| 5 | BFL | 0.002 | 0.944 | 1.000 | 0.005 | 0.876 | 1.000 | 0.023 | 0.931 | 1.000 | 0.004 | 0.888 | 1.000 |
| | BCL-OFS | 0.004 | 0.927 | 1.112 | 0.014 | 0.925 | 0.828 | 0.000 | 0.926 | 1.046 | 0.004 | 0.919 | 1.005 |
| 10 | BFL | 0.003 | 0.936 | 1.000 | 0.004 | 0.917 | 1.000 | 0.006 | 0.918 | 1.000 | 0.003 | 0.913 | 1.000 |
| | BCL-OFS | 0.010 | 0.945 | 0.987 | 0.006 | 0.944 | 0.912 | 0.005 | 0.973 | 0.768 | 0.009 | 0.958 | 0.882 |
| 20 | BFL | 0.004 | 0.959 | 1.000 | 0.002 | 0.931 | 1.000 | 0.013 | 0.952 | 1.000 | 0.003 | 0.931 | 1.000 |
| | BCL-OFS | 0.003 | 0.957 | 1.106 | 0.013 | 0.969 | 0.922 | 0.006 | 0.961 | 1.139 | 0.004 | 0.953 | 0.958 |
| 40 | BFL | 0.008 | 0.957 | 1.000 | 0.003 | 0.954 | 1.000 | 0.006 | 0.960 | 1.000 | 0.001 | 0.941 | 1.000 |
| | BCL-OFS | 0.003 | 0.950 | 1.006 | 0.005 | 0.949 | 1.113 | 0.002 | 0.964 | 1.038 | 0.004 | 0.950 | 0.968 |
| 100 | BFL | 0.005 | 0.954 | 1.000 | 0.000 | 0.948 | 1.000 | 0.002 | 0.965 | 1.000 | 0.003 | 0.963 | 1.000 |
| | BCL-OFS | 0.002 | 0.957 | 0.941 | 0.002 | 0.942 | 1.041 | 0.000 | 0.940 | 1.089 | 0.000 | 0.937 | 1.035 |

Web Table S6: Summary of $1,000$ simulations with $I = 5$, 10, 20, 40 and 100: bias, coverage probability (CP) and relative efficiency (RE) of pooled estimates of $AB$ and $AC$ treatment comparisons for two outcomes. The data generating mechanism was through an unequal between-study heterogeneity variance with ${\tau_1^{AB}}^2 = 0.25$, ${\tau_1^{AC}}^2 = 0.36$, ${\tau_2^{AB}}^2 = 0.36$, ${\tau_2^{AC}}^2 = 0.49$, $\mu_1^{AB} = 1$, $\mu_1^{AC} = 2$, $\mu_2^{AB} = -1$, and $\mu_2^{AC} = -2$, as well as the varying correlations. Results were based on the BFL method and the proposed BCL-OFS method.

| | | $\mu_1^{AB}$ | | | $\mu_1^{AC}$ | | | $\mu_2^{AB}$ | | | $\mu_2^{AC}$ | | |
|---|---|---|---|---|---|---|---|---|---|---|---|---|---|
| # Study | Method | Bias | CP | RE | Bias | CP | RE | Bias | CP | RE | Bias | CP | RE |
| 5 | BFL | 0.017 | 0.931 | 1.000 | 0.008 | 0.883 | 1.000 | 0.019 | 0.912 | 1.000 | 0.014 | 0.888 | 1.000 |
| | BCL-OFS | 0.003 | 0.941 | 0.911 | 0.009 | 0.927 | 0.880 | 0.022 | 0.929 | 0.945 | 0.008 | 0.927 | 0.881 |
| 10 | BFL | 0.007 | 0.931 | 1.000 | 0.010 | 0.889 | 1.000 | 0.003 | 0.930 | 1.000 | 0.010 | 0.901 | 1.000 |
| | BCL-OFS | 0.007 | 0.941 | 1.106 | 0.010 | 0.961 | 0.956 | 0.011 | 0.947 | 1.099 | 0.001 | 0.948 | 0.926 |
| 20 | BFL | 0.006 | 0.947 | 1.000 | 0.006 | 0.923 | 1.000 | 0.001 | 0.945 | 1.000 | 0.006 | 0.926 | 1.000 |
| | BCL-OFS | 0.004 | 0.957 | 1.077 | 0.005 | 0.961 | 1.037 | 0.002 | 0.953 | 0.997 | 0.001 | 0.960 | 0.870 |
| 40 | BFL | 0.006 | 0.957 | 1.000 | 0.001 | 0.944 | 1.000 | 0.004 | 0.965 | 1.000 | 0.005 | 0.942 | 1.000 |
| | BCL-OFS | 0.005 | 0.952 | 1.046 | 0.005 | 0.955 | 0.996 | 0.003 | 0.947 | 1.051 | 0.009 | 0.964 | 0.887 |
| 100 | BFL | 0.005 | 0.958 | 1.000 | 0.002 | 0.941 | 1.000 | 0.001 | 0.959 | 1.000 | 0.006 | 0.946 | 1.000 |
| | BCL-OFS | 0.002 | 0.949 | 1.043 | 0.002 | 0.941 | 1.003 | 0.001 | 0.931 | 1.069 | 0.000 | 0.920 | 0.966 |

Web Table S7: Comparisons of running time (in minutes) for the proposed method without and with the parallel sampling strategy (BCL/BCL Parallel), the Bayesian full-likelihood method with equal within-study correlations (BFL Equal Correlation), and the Bayesian full-likelihood method with unequal within-study correlations (BFL Unequal Correlation). The data is resampled from a real dataset.

| | | | | | |
|---|---|---|---|---|---|
| **Number of Treatment: 5** | | | | | |
| **Number of Studies** | **10** | **20** | **30** | **40** | **50** |
| BFL Unequal Correlation | 0.587 | 0.811 | 1.322 | 1.780 | 1.785 |
| BFL Equal Correlation | 0.310 | 0.474 | 0.753 | 1.041 | 1.064 |
| BCL | 0.478 | 0.616 | 0.833 | 1.031 | 1.075 |
| BCL Parallel | 0.192 | 0.221 | 0.268 | 0.318 | 0.314 |
| **Number of Treatment: 10** | | | | | |
| **Number of Studies** | **60** | **70** | **80** | **90** | **100** |
| BFL Unequal Correlation | 8.906 | 9.814 | 12.400 | 10.681 | 27.434 |
| BFL Equal Correlation | 3.248 | 3.714 | 4.536 | 4.296 | 5.319 |
| BCL | 1.495 | 1.632 | 1.911 | 1.798 | 1.969 |
| BCL Parallel | 0.767 | 0.835 | 0.977 | 0.918 | 1.005 |
| **Number of Treatment: 20** | | | | | |
| **Number of Studies** | **110** | **120** | **130** | **140** | **150** |
| BFL Unequal Correlation | 59.520 | 66.619 | 71.195 | 76.159 | 78.046 |
| BFL Equal Correlation | 13.065 | 14.540 | 15.581 | 16.801 | 17.351 |
| BCL | 4.882 | 5.167 | 5.700 | 6.075 | 6.199 |
| BCL Parallel | 2.427 | 2.613 | 2.845 | 3.034 | 3.105 |

#### Appendix E: MCMC setup for real applications

This section revisited the two motivating NMA datasets introduced in Section 2 of the main text. While the original NMA analyses likely incorporated known within-study correlations in their models, these correlations were not explicitly reported in the raw datasets. We addressed this limitation by implementing the proposed method on both datasets without requiring knowledge of within-study correlations. For both re-analyses, we applied non-informative prior distributions and estimated relative treatment comparisons using odds ratios (OR) and risk ratios (RR). Treatment comparisons were ranked using the surface under the cumulative ranking curve (SUCRA) (Salanti et al., 2011) for each outcome. The MCMC setup involved generating five independent chains of $50,000$ iterations each for every parameter. The initial $10,000$ iterations were discarded as burn-in, and posterior samples were collected every 5th iteration thereafter. Convergence of Markov chain simulation was assessed via the traceplots and the potential scale reduction factor (PSRF) (Gelman et al., 2014), with values below 1.05 indicating satisfactory convergence.

#### Appendix F: Potential scale reduction factor

To diagnose convergence in the Markov chain Monte Carlo (MCMC) simulations, we utilized chains with different initial values. After discarding the first half as warm-up, we have $L$ steps in each chain. Next, we split each chain into two parts. This resulted in $m = 10$ chains, each containing $n = L/2$ steps. We then calculated the between- and within-sequence variances to evaluate convergence.

$$B = \frac{n}{m-1} \sum_{j=1}^{m} \left( \bar{\psi}_{.j} - \bar{\psi}_{..} \right)^2, \quad \text{where } \bar{\psi}_{.j} = \frac{1}{n} \sum_{i=1}^{n} \psi_{ij}, \bar{\psi}_{..} = \frac{1}{m} \sum_{j=1}^{m} \bar{\psi}_{.j};$$

$$W = \frac{1}{m} \sum_{j=1}^{m} s_j^2, \quad \text{where } s_j^2 = \frac{1}{n-1} \sum_{i=1}^{n} \left( \psi_{ij} - \bar{\psi}_{.j} \right)^2.$$

The marginal posterior variance is calculated based on the weighted average of $B$ and $W$

$$\widehat{\text{var}}^{+}(\psi \mid y) = \frac{n-1}{n} W + \frac{1}{n} B.$$

Given this quantity overestimates the marginal posterior variance, we calculate the potential scale reduction index (Gelman et al., 2014)

$$\hat{R} = \sqrt{\frac{\widehat{\text{var}}^{+}(\psi \mid y)}{W}}.$$

We determine whether the posterior sample converges convergences by checking if this factor is below 1.2.

### Appendix G: Additional results for real application

Web Table S8: Potential scale reduction factor for the root coverage data

| | CRC | RecRed | CAL gain | KT gain |
|---|---|---|---|---|
| CAF+EMD | 1.00 | 1.00 | 1.00 | 1.00 |
| CAF+ADM | 1.00 | 1.00 | 1.00 | 1.00 |
| CAF+PRP | 1.00 | 1.00 | 1.00 | 1.00 |
| CAF+HF-DDS | 1.00 | | | |
| CAF+CM | 1.00 | 1.00 | 1.00 | 1.00 |
| CAF | 1.00 | 1.00 | 1.00 | 1.00 |
| CAF+CTG | 1.00 | 1.00 | 1.00 | 1.00 |

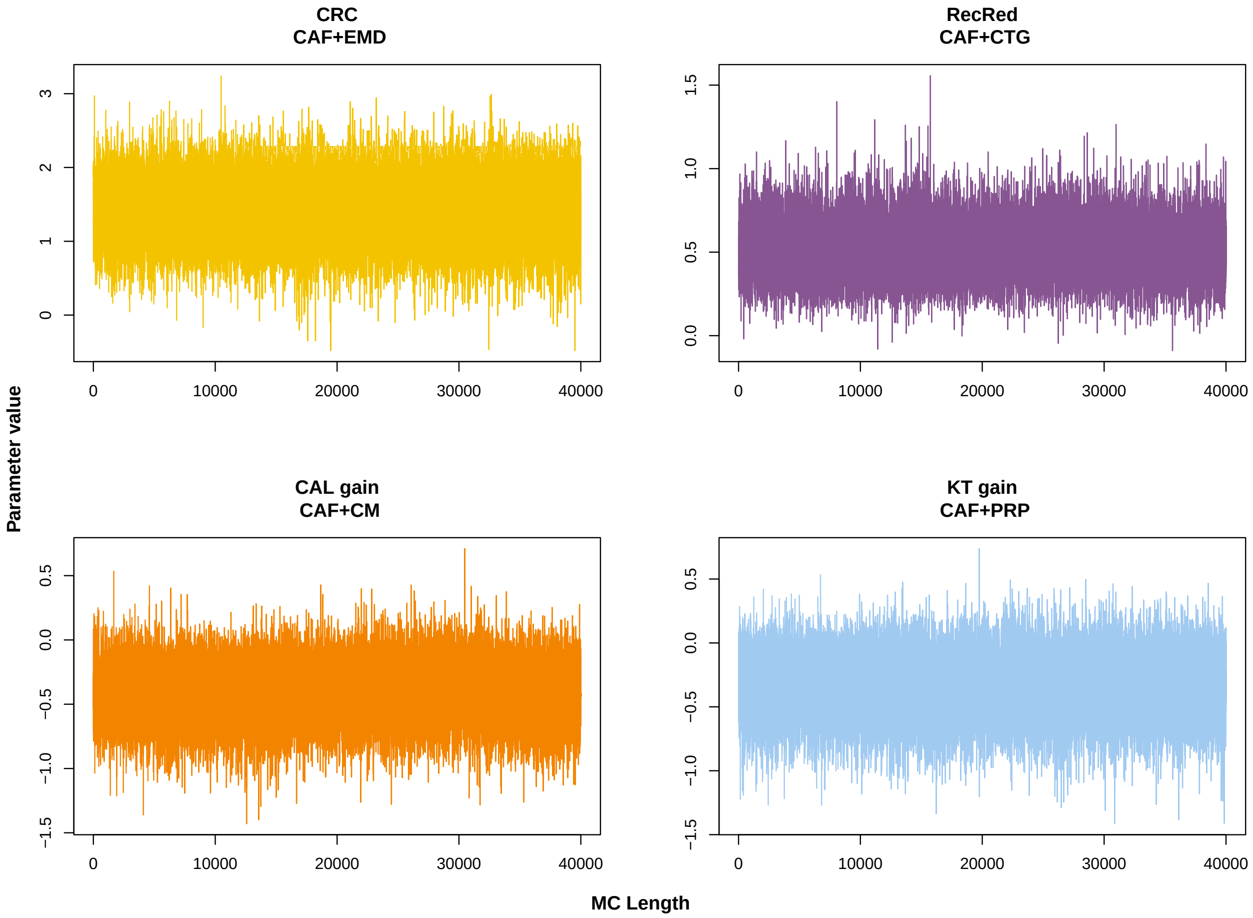


Web Figure S4: Trace plot for randomly selected pooled estimates of treatment effects for the four outcomes in the the root coverage data.

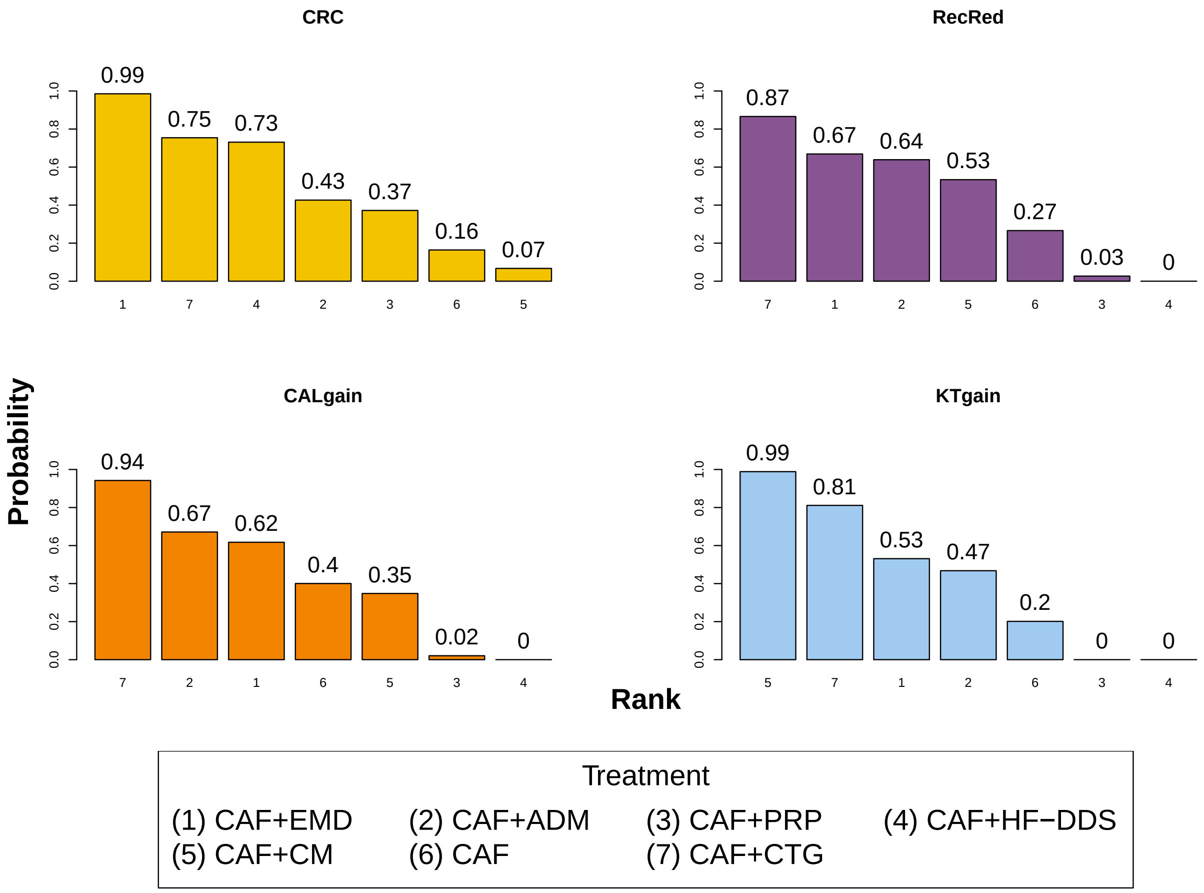


Web Figure S5: Treatment ranking was determined by integrating posterior estimates derived from the BCL-OFS method into the surface under the cumulative ranking method for the root coverage data.

| | CAF+EMD | CAF+ADM | CAF+PRP | CAF+HF-DDS | CAF+CM | CAF | CAF+CTG | CAF+BM |
|---|---|---|---|---|---|---|---|---|
| CAF+EMD | CAF+EMD | 0.42 (0.18, 0.83) | 0.31 (0.19, 0.47) | 0.63 (0.40, 0.94) | 0.17 (0.05, 0.41) | 0.25 (0.15, 0.38) | 0.63 (0.40, 0.93) | 0.26 (0.13, 0.46) |
| CAF+ADM | 0.41 (0.13, 1.38) | CAF+ADM | 0.91 (0.35, 1.72) | 1.71 (0.95, 2.67) | 0.47 (0.12, 1.13) | 0.73 (0.28, 1.38) | 1.69 (0.95, 2.65) | 0.72 (0.30, 1.36) |
| CAF+PRP | 0.33 (0.05, 2.24) | 0.81 (0.11, 5.91) | CAF+PRP | 2.09 (1.37, 3.11) | 0.57 (0.16, 1.34) | 0.80 (0.79, 0.81) | 2.07 (1.36, 3.09) | 0.86 (0.46, 1.45) |
| CAF+HF-DDS | 0.64 (0.02, 22.58) | 1.58 (0.04, 53.89) | 1.94 (0.03, 97.71) | CAF+HF-DDS | 0.28 (0.09, 0.62) | 0.41 (0.26, 0.59) | 0.99 (0.92, 1.02) | 0.42 (0.23, 0.68) |
| CAF+CM | 0.14 (0.02, 1.08) | 0.35 (0.04, 2.51) | 0.43 (0.03, 5.57) | 0.22 (0.00, 10.30) | CAF+CM | 2.11 (0.60, 4.90) | 5.13 (1.61, 11.48) | 2.17 (0.57, 5.24) |
| CAF | 0.26 (0.11, 0.57) | 0.63 (0.22, 1.56) | 0.78 (0.14, 4.17) | 0.40 (0.01, 16.67) | 1.82 (0.26, 14.29) | CAF | 2.59 (1.70, 3.85) | 1.07 (0.57, 1.81) |
| CAF+CTG | 0.61 (0.24, 1.52) | 1.49 (0.55, 3.45) | 1.82 (0.28, 12.50) | 0.94 (0.03, 33.33) | 4.35 (0.75, 25.00) | 2.37 (1.18, 5.09) | CAF+CTG | 0.42 (0.23, 0.68) |
| CAF+BM | 0.24 (0.08, 0.72) | 0.60 (0.18, 1.75) | 0.74 (0.11, 5.00) | 0.38 (0.01, 14.29) | 1.72 (0.25, 12.50) | 0.96 (0.39, 2.31) | 0.40 (0.18, 0.84) | CAF+BM |

Web Figure S6: Network meta-analysis of the outcome of CRC in root coverage data. Results are the odds ratios, with the column-defining treatment compared to the row-defining treatment. The lower triangular matrix refers to the standard Bayesian NMA using Lu and Ades' approach, and the upper triangular matrix refers to the BCL-OFS method.

| | CAF+EMD | CAF+ADM | CAF+PRP | CAF+CM | CAF | CAF+CTG | CAF+BM |
|---|---|---|---|---|---|---|---|
| CAF+EMD | CAF+EMD | 0.01 (-0.50, 0.59) | -0.62 (-0.97, -0.30) | 0.00 (-0.80, 1.15) | -0.41 (-0.76, -0.09) | 0.22 (-0.27, 0.84) | -0.29 (-0.77, 0.29) |
| CAF+ADM | 0.14 (-0.66, 1.12) | CAF+ADM | -0.62 (-1.14, -0.21) | -0.01 (-0.66, 0.93) | -0.42 (-0.92, -0.01) | 0.22 (-0.16, 0.64) | -0.29 (-0.70, 0.14) |
| CAF+PRP | -0.49 (-1.76, 0.89) | -0.63 (-1.93, 0.65) | CAF+PRP | 0.61 (-0.17, 1.80) | 0.20 (0.18, 0.24) | 0.84 (0.43, 1.40) | 0.33 (-0.05, 0.85) |
| CAF+CM | 0.10 (-1.01, 1.39) | -0.04 (-1.17, 1.10) | 0.59 (-0.96, 2.17) | CAF+CM | -0.41 (-1.58, 0.36) | 0.22 (-0.49, 0.69) | -0.28 (-1.09, 0.26) |
| CAF | -0.32 (-0.92, 0.43) | -0.46 (-1.16, 0.17) | 0.17 (-0.96, 1.28) | -0.42 (-1.54, 0.64) | CAF | 0.64 (0.24, 1.19) | 0.13 (-0.25, 0.63) |
| CAF+CTG | 0.36 (-0.33, 1.19) | 0.22 (-0.42, 0.86) | 0.85 (-0.40, 2.10) | 0.26 (-0.69, 1.17) | 0.68 (0.15, 1.25) | CAF+CTG | -0.51 (-0.75, -0.29) |
| CAF+BM | -0.24 (-1.05, 0.69) | -0.38 (-1.25, 0.40) | 0.25 (-1.07, 1.53) | -0.34 (-1.48, 0.75) | 0.08 (-0.58, 0.72) | -0.59 (-1.25, -0.03) | CAF+BM |

Web Figure S7: Network meta-analysis of the outcome of RecRed in root coverage data. The results are the standard mean difference, with the column-defining treatment compared to the row-defining treatment. The lower triangular matrix refers to the standard Bayesian NMA using Lu and Ades' approach, and the upper triangular matrix refers to to the BCL-OFS method.

| | CAF+EMD | CAF+ADM | CAF+PRP | CAF+CM | CAF | CAF+CTG | CAF+BM |
|---|---|---|---|---|---|---|---|
| CAF+EMD | CAF+EMD | -0.07 (-0.36, 0.24) | -0.70 (-0.89, -0.52) | 1.02 (0.61, 1.42) | -0.40 (-0.59, -0.22) | 0.79 (0.46, 1.12) | -0.37 (-0.77, 0.05) |
| CAF+ADM | -0.03 (-0.73, 0.72) | CAF+ADM | -0.64 (-0.88, -0.41) | 1.09 (0.68, 1.47) | -0.34 (-0.58, -0.11) | 0.86 (0.54, 1.16) | -0.30 (-0.75, 0.15) |
| CAF+PRP | -0.65 (-1.84, 0.55) | -0.62 (-1.83, 0.59) | CAF+PRP | 1.72 (1.34, 2.09) | 0.30 (0.29, 0.31) | 1.49 (1.20, 1.78) | 0.33 (-0.02, 0.70) |
| CAF+CM | 1.05 (0.05, 2.07) | 1.08 (0.10, 2.04) | 1.70 (0.27, 3.12) | CAF+CM | -1.42 (-1.79, -1.05) | -0.23 (-0.46, 0.01) | -1.39 (-1.86, -0.89) |
| CAF | -0.36 (-0.86, 0.17) | -0.34 (-0.89, 0.21) | 0.29 (-0.79, 1.37) | -1.41 (-2.33, -0.48) | CAF | 1.19 (0.90, 1.49) | 0.03 (-0.32, 0.40) |
| CAF+CTG | 0.82 (-0.20, 1.46) | 0.85 (0.29, 1.40) | 1.47 (0.29, 2.65) | -0.23 (-1.01, 0.57) | 1.18 (0.70, 1.66) | CAF+CTG | -1.16 (-1.57, -0.73) |
| CAF+BM | -0.31 (-1.01, 0.44) | -0.28 (-0.96, 0.41) | 0.34 (-0.87, 1.56) | -1.36 (-2.28, -0.40) | 0.05 (-0.51, 62.00) | -1.13 (-1.62, -0.63) | CAF+BM |

Web Figure S8: Network meta-analysis of the outcome of KT gain in root coverage data. The results are the standard mean difference, with the column-defining treatment compared to the row-defining treatment. The lower triangular matrix refers to the standard Bayesian NMA using Lu and Ades' approach, and the upper triangular matrix refers to the BCL-OFS method.

| | CAF+EMD | CAF+ADM | CAF+PRP | CAF+CM | CAF | CAF+CTG | CAF+BM |
|---|---|---|---|---|---|---|---|
| CAF+EMD | CAF+EMD | 0.06 (-0.76, 0.88) | -0.71 (-1.23, -0.19) | -0.26 (-1.20, 0.68) | -0.21 (-0.73, 0.31) | 0.43 (-0.49, 1.38) | 0.15 (-0.73, 1.06) |
| CAF+ADM | 0.76 (-1.25, 2.85) | CAF+ADM | -0.77 (-1.20, -0.34) | -0.32 (-0.73, 0.09) | -0.27 (-0.7, 0.16) | 0.37 (0.00, 0.78) | 0.09 (-0.34, 0.54) |
| CAF+PRP | -0.15 (-3.06, 2.64) | -0.92 (-3.77, 1.80) | CAF+PRP | 0.44 (-0.10, 1.01) | 0.50 (0.48, 0.52) | 1.14 (0.62, 1.70) | 0.86 (0.33, 1.42) |
| CAF+CM | 0.56 (-2.04, 3.27) | -0.20 (-2.83, 2.54) | 0.72 (-2.66, 4.16) | CAF+CM | 0.06 (-0.50, 0.60) | 0.70 (0.54, 0.87) | 0.42 (0.09, 0.74) |
| CAF | 0.41 (-1.11, 2.02) | -0.35 (-1.87, 1.24) | 0.56 (-1.61, 2.87) | -0.15 (-2.77, 2.42) | CAF | 0.64 (0.12, 1.20) | 0.36 (-0.17, 0.92) |
| CAF+CTG | 1.14 (-0.42, 2.78) | 0.38 (-1.32, 1.90) | 1.30 (-1.25, 3.98) | 0.58 (-1.70, 2.73) | 0.74 (-0.60, 2.03) | CAF+CTG | -0.28 (-0.58, 0.00) |
| CAF+BM | 0.64 (-1.51, 2.88) | -0.12 (-2.32, 1.95) | 0.80 (-1.82, 3.45) | 0.08 (-2.57, 2.62) | 0.23 (-1.52, 1.90) | -0.50 (-2.04, 1.16) | CAF+BM |

Web Figure S9: Network meta-analysis of the outcome of CAL gain in root coverage data. Results are the standard mean difference, with the column-defining treatment compared to the row-defining treatment. The lower triangular matrix refers to the standard Bayesian NMA using Lu and Ades' approach, and the upper triangular matrix refers to the BCL-OFS method.

Web Table S9: Potential scale reduction factor for the first part of the anaemia data

| | Blood transfusion | Cardiovascular death | Death (any cause) | Hypertension | Kidney failure |
|---|---|---|---|---|---|
| Biosimilar darbepoetin alfa | 1.00 | | 1.00 | 1,00 | 1.00 |
| Biosimilar epoetin | 1.00 | 1.00 | 1.00 | 1.00 | |
| Darbepoetin alfa | 1.00 | 1.00 | 1.00 | 1.00 | 1.00 |
| Epoetin beta | 1.00 | 1.00 | 1.00 | 1.00 | 1.00 |
| Methoxy polyethylene glycol-epoetin beta | 1.00 | 1.00 | 1.00 | 1.00 | 1.00 |
| Epoetin alpha | 1.00 | 1.00 | 1.00 | 1.00 | 1.00 |

Web Table S10: Potential scale reduction factor for the second part of the anaemia data

| | Major cardiovascular events | Myocardial infarction | Stroke | Vascular access thrombosis |
|---|---|---|---|---|
| Biosimilar darbepoetin alfa | | 1.00 | 1.00 | 1.00 |
| Biosimilar epoetin | 1.00 | 1.00 | 1.00 | 1.00 |
| Darbepoetin alfa | 1.00 | 1.00 | 1.00 | 1.00 |
| Epoetin beta | 1.00 | 1.00 | 1.00 | 1.00 |
| Methoxy polyethylene glycol-epoetin beta | 1.00 | 1.00 | 1.00 | 1.00 |
| Epoetin alpha | 1.00 | 1.00 | 1.00 | 1.00 |

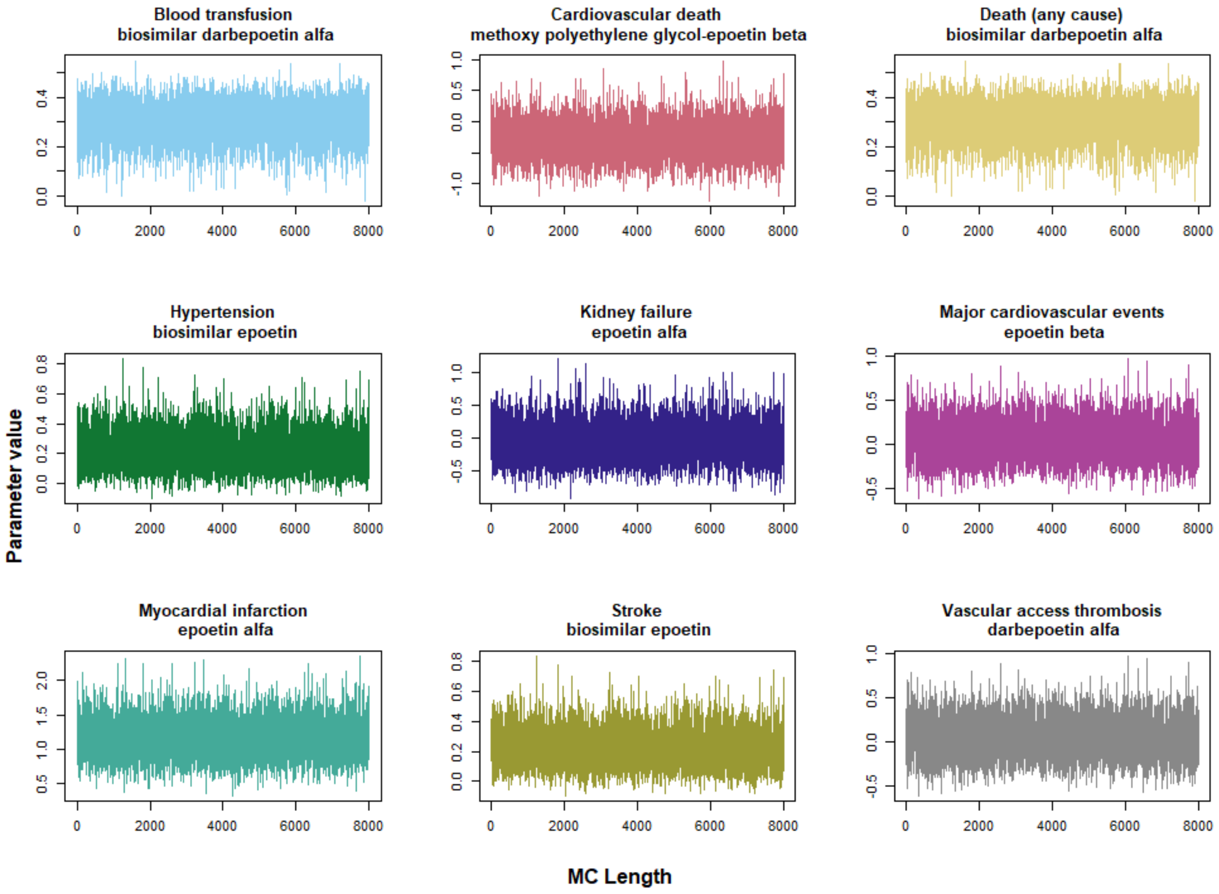


Web Figure S10: Trace plot for randomly selected pooled estimates of treatment effects for the nine outcomes in the the anaemia data.

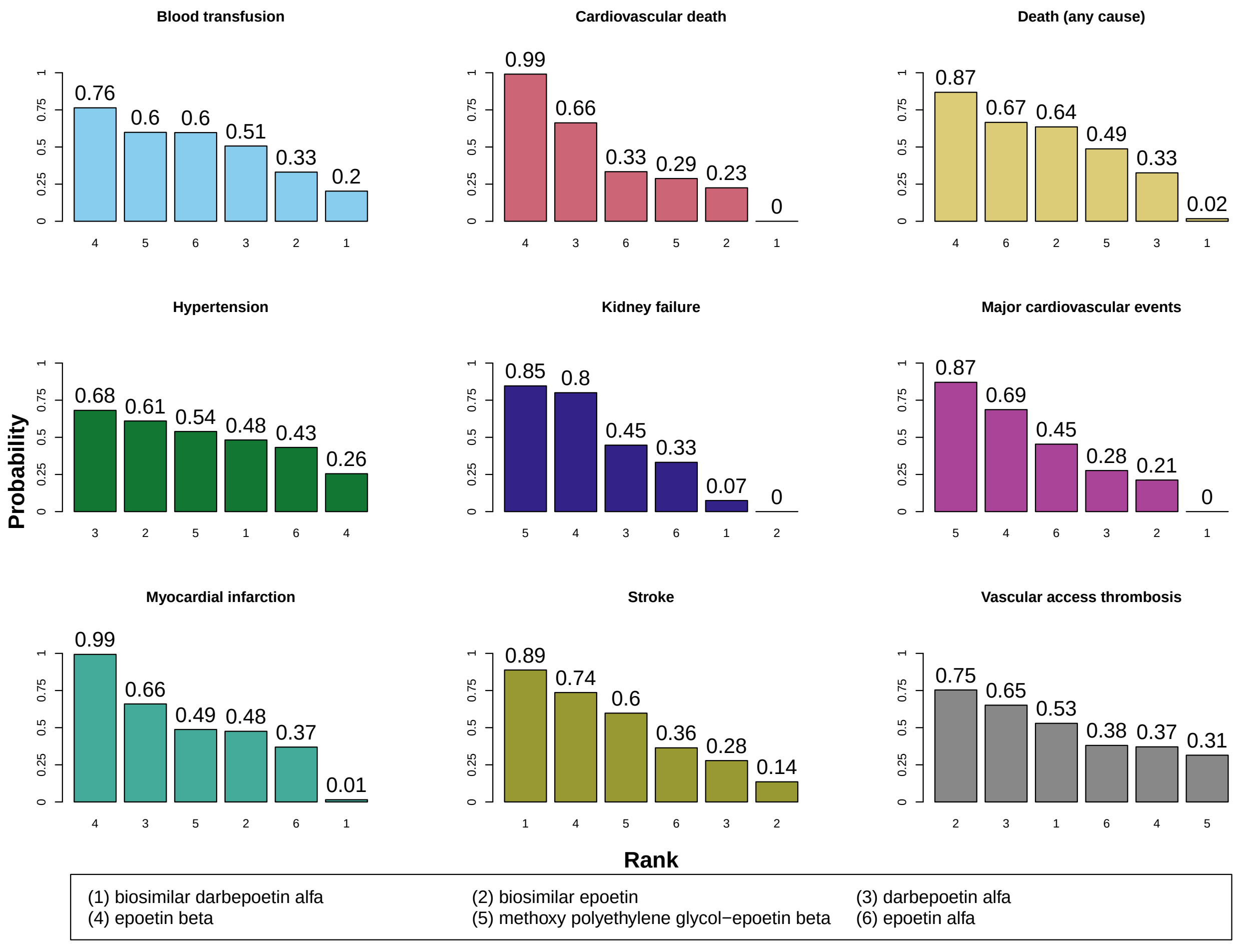


Web Figure S11: Treatment ranking was determined by integrating posterior estimates derived from the BCL-OFS method into the surface under the cumulative ranking method for the anaemia data. Placebo was considered the reference treatment.

| | Biosimilar darbepoetin alfa | Biosimilar epoetin | Darbepoetin alfa | Epoetin beta | Methoxy polyethylene glycol-epoetin beta | Epoetin alfa | Placebo |
|---|---|---|---|---|---|---|---|
| Biosimilar darbepoetin alfa | Biosimilar darbepoetin alfa | 1.53 (0.20, 5.58) | 1.77 (0.26, 6.06) | 2.68 (0.31, 10.06) | 2.10 (0.24, 7.97) | 1.74 (0.32, 5.63) | 0.59 (0.08, 2.07) |
| Biosimilar epoetin | 1.10 (0.21, 5.91) | Biosimilar epoetin | 1.41 (0.37, 3.77) | 2.09 (0.44, 6.32) | 1.66 (0.33, 5.13) | 1.35 (0.55, 2.92) | 0.46 (0.12, 1.20) |
| Darbepoetin alfa | 1.36 (0.27, 6.83) | 1.23 (0.38, 3.96) | Darbepoetin alfa | 1.63 (0.47, 4.17) | 1.00 (0.39, 2.19) | 1.16 (0.44, 2.68) | 0.37 (0.13, 0.82) |
| Epoetin beta | 1.94 (0.32, 11.59) | 1.76 (0.45, 6.79) | 1.43 (0.47, 4.34) | Epoetin beta | 1.51 (0.41, 4.03) | 0.88 (0.25, 2.31) | 0.25 (0.10, 0.52) |
| Methoxy polyethylene glycol-epoetin beta | 1.13 (0.19, 6.75) | 1.02 (0.25, 4.09) | 1.21 (0.50, 2.92) | 1.72 (0.55, 5.43) | Methoxy polyethylene glycol-epoetin beta | 1.15 (0.30, 3.26) | 0.35 (0.10, 0.91) |
| Epoetin alfa | 1.34 (0.31, 5.81) | 1.21 (0.53, 2.76) | 0.98 (0.41, 2.37) | 0.69 (0.23, 2.06) | 1.19 (0.38, 3.75) | Epoetin alfa | 0.35 (0.12, 0.73) |
| Placebo | 0.37 (0.07, 1.91) | 0.34 (0.11, 1.03) | 0.27 (0.11, 0.69) | 0.19 (0.08, 0.47) | 0.33 (0.11, 1.02) | 0.28 (0.13, 0.61) | Placebo |

Web Figure S12: Network meta-analysis of the outcome of preventing blood transfusion in CKD patients with anaemia. Results are the odds ratios, with the column-defining treatment compared to the row-defining treatment. The lower triangular matrix refers to the standard NMA using White's approach, and the upper triangular matrix refers to the BCL-OFS method.

| | Biosimilar darbepoetin alfa | Biosimilar epoetin | Darbepoetin alfa | Epoetin beta | Methoxy polyethylene glycol-epoetin beta | Epoetin alfa | Placebo |
|---|---|---|---|---|---|---|---|
| Biosimilar darbepoetin alfa | Biosimilar darbepoetin alfa | 2.38 (0.59, 6.66) | 1.92 (0.55, 4.95) | 2.71 (0.68, 7.47) | 2.12 (0.54, 5.91) | 2.37 (0.61, 6.44) | 1.92 (0.54, 5.01) |
| Biosimilar epoetin | 2.04 (0.58, 7.17) | Biosimilar epoetin | 0.87 (0.50, 1.42) | 1.22 (0.61, 2.23) | 0.96 (0.46, 1.79) | 1.03 (0.72, 1.42) | 0.86 (0.49, 1.42) |
| Darbepoetin alfa | 1.64 (0.52, 5.18) | 0.80 (0.48, 1.34) | Darbepoetin alfa | 1.41 (0.87, 2.15) | 0.95 (0.60, 1.44) | 1.23 (0.80, 1.80) | 1.00 (0.77, 1.22) |
| Epoetin beta | 2.35 (0.66, 8.32) | 1.15 (0.59, 2.24) | 1.43 (0.84, 2.44) | Epoetin beta | 1.33 (0.69, 2.36) | 0.92 (0.49, 1.54) | 0.74 (0.47, 1.11) |
| Methoxy polyethylene glycol-epoetin beta | 1.52 (0.45, 5.18) | 0.75 (0.39, 1.44) | 1.08 (0.71, 1.65) | 1.54 (0.80, 2.97) | Methoxy polyethylene glycol-epoetin beta | 1.17 (0.61, 2.02) | 0.95 (0.56, 1.50) |
| Epoetin alfa | 2.07 (0.61, 6.97) | 1.01 (0.74, 1.39) | 1.26 (0.84, 1.89) | 0.88 (0.48, 1.61) | 1.36 (0.76, 2.43) | Epoetin alfa | 0.84 (0.54, 1.25) |
| Placebo | 1.63 (0.51, 5.23) | 0.80 (0.47, 1.36) | 0.99 (0.81, 1.21) | 0.69 (0.40, 1.20) | 1.07 (0.67, 1.71) | 0.79 (0.51, 1.22) | Placebo |

Web Figure S13: Network meta-analysis of the outcome of preventing all-cause death in CKD patients with anaemia. Results are the odds ratios, with the column-defining treatment compared to the row-defining treatment. The lower triangular matrix refers to the standard NMA using White's approach, and the upper triangular matrix refers to the BCL-OFS method.
